# Global evolutionary epidemiology, phylogeography and resistome dynamics of *Citrobacter species, Enterobacter hormaechei, Klebsiella variicola, and Proteeae clones: A One Health analyses*

**DOI:** 10.1101/2020.05.21.20109504

**Authors:** John Osei Sekyere, Melese Abate Reta

## Abstract

**Background.:** The global epidemiology and resistomes dynamics of multidrug-resistant *Citrobacter spp., Enterobacter hormaechei, Klebsiella variicola, morganella morganii, Proteus mirabilis* and *Providencia spp*. have not been described, despite their importance as emerging opportunistic clinical pathogens.

**Methods.:** The genomes of the above-mentioned organisms were curated from PATRIC and NCBI and used for evolutionary epidemiology, phylogeography and resistome analyses. The phylogeny trees were drawn using RAXmL and edited with Figtree. The resistomes were curated from GenBank and the phylogeography was manually mapped.

**Results and conclusion.:** *Mcr-9* and other *mcr* variants were highly prevalent in *E. hormaechei subsp*. and substantial in *C. freundii* whilst KPC, OXA-48, NDM, IMP, VIM, TEM, OXA and SHV were abundant in global *E. hormaechei subsp., Citrobacter freundii, P. mirabilis, P. stuartii* and *P. rettgeri* clones/clades. Species-specific ampCs were highly conserved in respective species whilst fluoroquinolones, aminoglycosides, macrolides, fosfomycin, chloramphenicol, tetracycline, sulphamethoxazole and trimethoprim resistance mechanisms were abundantly enriched in almost all clades of most of the species, making them extensively and pandrug resistant; *K. variicola, C. amalonaticus* and *C, koseri* had relatively few resistance genes. Vertical and horizontal resistome transmissions as well as local and international dissemination of strains evolving from common ancestors were observed, suggesting the anthroponotic, zoonotic, and food-/water-borne infectiousness of these pathogens. There is a global risk of pandrug resistant strains escalating local and international outbreaks of antibiotic-insensitive infections, initiating the dawn of a post-antibiotic era.

## Introduction

Antibiotic resistance is mainly disseminated via horizontal and vertical transmission through mobile genetic elements such as plasmids and transposons and through clonal and multiclonal expansion of same species ^1–6^. Conjugative plasmids have been implicated in the transmission of several resistance determinants within and across species, resulting in the presence of same or very similar resistomes in same and different species and clones ^1,4,7–9^. Thus, the emergence of plasmid-borne resistance genes is always a cause for concern as they help breach the species barrier and shuttle resistance genes (ARGs) from commensals and non-pathogenic bacteria to pathogenic ones or vice versa ^3,4,10,11^. Such has been the case with the emergence and rapid spread of extended-spectrum β-lactamases (ESBLs) viz., TEM, SHV, OXA and CTX-M, carbapenemases such as NDM, IMP, VIM, KPC and GES, the mobile colistin resistance gene *mcr-1* (to *mcr-10*) and recently, the mobile tigecycline resistance gene, *tet*(*X*) ^12–17^ Thus, such conjugative plasmids influence the genomic plasticity of several related and unrelated species and genera of bacteria ^8,11,18–21^.

Coupled with plasmid-borne dissemination of ARGs is the selection and expansion of specific drug-resistant clones ^11,21^, which quickly spread under antibiotic pressure to overpopulate their environments, facilitating their survival and subsequent spread to other environments ^14,22,23^. In cases where such clones harbour resistance plasmids, their expansion almost always lead to the concomitant replication and intra-clonal as well as inter-clonal spread of such plasmids ^4,5,7,8,24,25^ Thus, as such clones are disseminated through contact, food, water, farms, hospitals, and the environment, they carry with them these resistance plasmids to colonize new hosts and environments ^6,26–28^. It is thus not surprising to have same clones hosting the same plasmids, contain the same resistomes ^4–6,21,28^. This explains the presence of multi-drug resistance (MDR) in particular international clones such as *Klebsiella pneumoniae* ST258 and *E. coli* ST113 ^21,25,29,30^.

Hence, tracing the phylogeography of clones and their associated resistance genes is highly critical in epidemiology and public health as it provides necessary data to contain the further spread of ARGs ^26,28,31^. In this work, the global evolutionary epidemiology and resistome dynamics of clinically important but relatively less isolated Enterobacteriaceae pathogens are described ^8^. It is notable that most of the recently emerged or novel resistance genes in bacteria have occurred in Enterobacteriaceae more than in any other family of bacteria, making Enterobacteriaceae particularly important medically ^28,32,33^. These include ESBL-, carbapenemase-, *mcr-* and *tet*(*X*)*-producing* producing Enterobacteriaceae, which have been classified by the WHO as high and critical priority pathogens due to their implication in high mortalities and morbidities ^32,33^. Although *Citrobacter spp., Enterobacter hormaechei, Klebsiella variicola, Morganella morganii, Proteus spp*. and *Providencia spp*. are not mostly reported as *Escherichia coli, Klebsiella pneumoniae*, and *Salmonella enterica*, they have been associated with multiple resistance and clinical fatalities ^3,7,14,25,28,34^ Due to the transferability of resistance plasmids between members of the Enterobacteriaceae, the global resistome epidemiology of these six genera is important as they could be eventually transferred to commonly isolated Enterobacteriaceae species ^11,14,21,25,35,36^.

## Results

### Included genomes

A total of 2,377 genomes from *C. freundii* (n=569 genomes), *C. koseri* (n=82 genomes), *C. amalonaticus* (n=35 genomes), *E. steigerwaltii* and *E oharae* (n=121 genomes), *E. xiangfangensis* (n= 90 genomes), *E. hormaechei* (n=563 genomes), *K. variicola* (n=574 genomes), *M. morganii* (n=59 genomes), *P. mirabilis* (n= 156 genomes), and *Providencia spp*. (n=128 genomes) were obtained from PATRIC and NCBI databases as at January 2020 and used for downstream analyses (Tables S1-S3). Included in *C. freundii, C. koseri, E. xiangfangesis, K. variicola* and *Providencia spp*. genomes were genomes of other Gram-negative bacterial species or genera that were initially classified within these respective species but later reclassified by NCBI’s ANI (average nucleotide identity) analysis. The genomes of these reclassified species were however maintained and included in the resistome analyses to serve as controls for comparison (Tables S1-S3).

The genomes were mainly isolated from human specimens, followed by animal (including food animals), plants (including food crops) and environmental specimens. The human and animal specimens used included urine, blood, stool, catheter tip, swabs etc. whilst the environmental specimens used included soils, hospital environments, water, wastewater, sinks etc. (Tables S1-S3). In all, these genomes were obtained from 67 countries globally, with the USA having the most genomes for all species: *C. freundii/spp*. (USA=233, China=34, France=14, Spain=11); *C. amalonaticus* (USA=17); *C. koseri* (USA=55); *E. steigerwaltii/oharae* (USA=40, Japan=15); *E. hormaechei* (USA=142, China=78, Japan=72, France=13, UK=10); *E. xiangfangensis* (USA=50, China=14, India=12); *K. variicola* (USA=289, Germany=50, China=24, Bangladesh=23); *M. morganii* (USA=16); *P. mirabilis* (USA=38, France=27, China=17); *Providencia spp*. (USA=61).

*C. freundii*, the only species among the species included in this analysis to have an MLST scheme, had 84 different clones or sequence types (STs). ST100 (n=51), ST22 (n=51), ST62 (n=18), ST11 (n=14), ST299 (n=11), ST8 (n=10), ST114 (n=8), and ST98 (n=8) were the commonest clones.

### Species epidemiology

#### Citrobacter species

Amongst the *C. freundii* genomes were other *Citrobacter spp*. such as *C. werkmanii, C. youngae, C. brakii, C. portucalensis* etc. *C. werkmanii* were isolated from humans, sprouts, and sinks whilst *C. brakii* were obtained from humans, vegetables, and hospital environments. Some C. *werkmanii* and *C. brakii* strains were found in USA, Germany, and India (Fig. S1A-B). *C. koseri* were mainly found from humans and from a mouse (clade B2) (Fig. S1A-B). A *Citrobacter spp*. (clade A) outbreak was observed in the US (Texas, Boston) and Mexico (Fig. S1A-B). *C. portucalensis* of the same clade (but different clones) were found in Nigeria (vegetables) and Brazil (turtle) as well as from effluents (UK), humans (China), chives, carrots and salad (Germany); these clustered in *C. freundii* clade B1. *C. freundii* strains of different STs and countries clustered together into clades, showing the wide distribution of strains from the same ancestor (or of close evolutionary distance); they were isolated from humans, plants, animals, and the environment (Fig. 1 & S1A-B).

**Figure 1.**
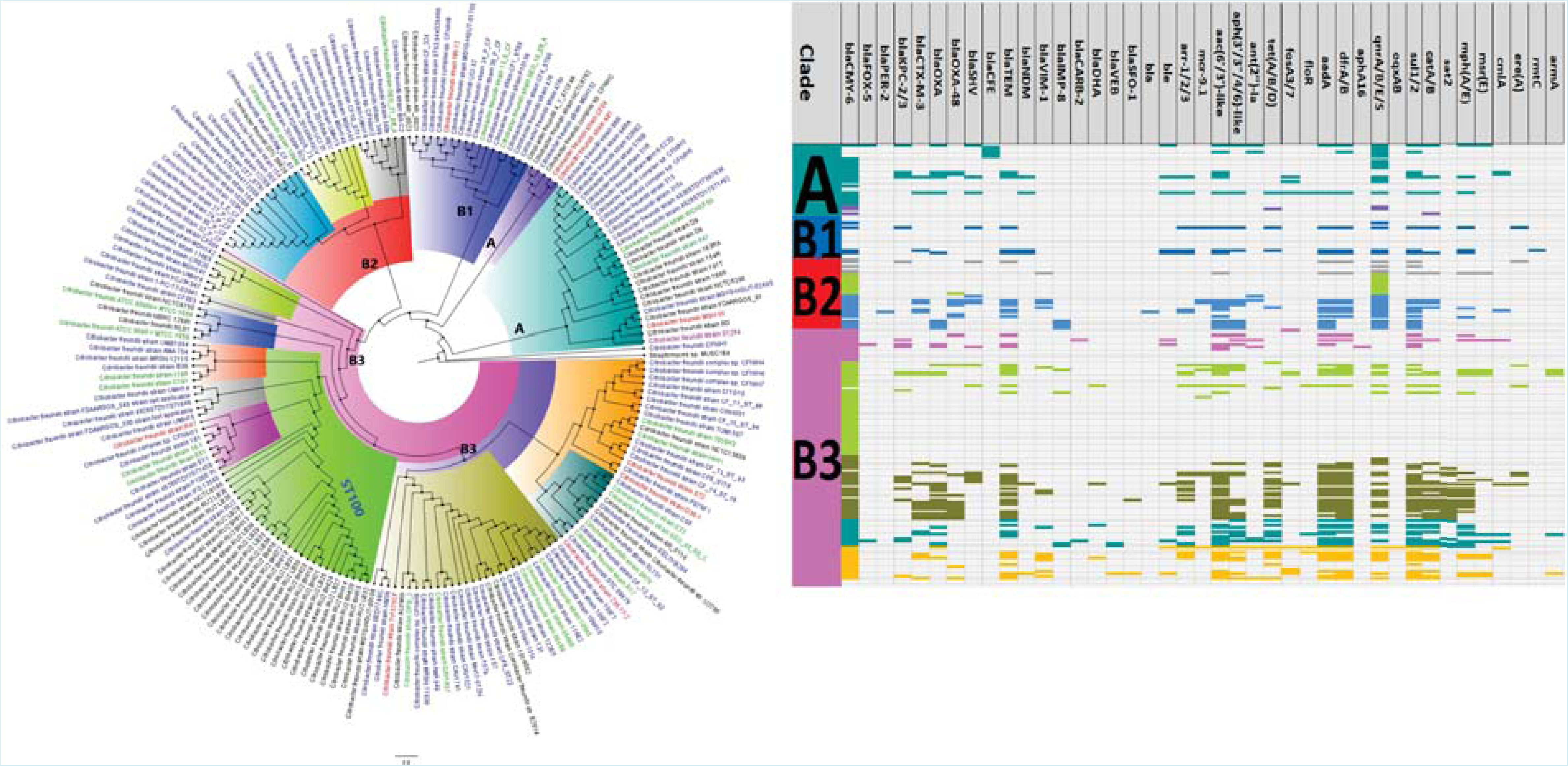
Evolutionary epidemiology and resistome of global *Citrobacter freundii* isolates. *C. freundii* clustered into four main clades (A, B1, B2 and B3), highlighted with distinct colours. Clade B3 had the most resistome abundance and diversity. Strains from humans (blue labels), animals (red labels), plants (purple/mauve labels) and the environment (green labels) were found in the same clade/cluster. *Bla*_CMY_ was conserved in these genomes.

*C. werkmannii, C. brakii* and *C. youngae* clustered within *C. freundii* clade A whilst *C. portucalensis* and *C. youngae*, were clustered in clade B; *C. koseri* clustered in clade B3 of *C. freundii* (Fig. 1 & S1A-B). *C. freundii* clades B2 and B3 had a richer resistome than clades A and B1, although bla_CMY_, *qnr, sul1/2*, aac(6’/3’)-like ARGs were common in all the clades; mph(A/E), *catAB, dfrAB, aadA* and bla_TEM_ were common in clades B2 and B3. Important ARGs such as *mcr*, bla_KPc_, bla_CTX-M_, bla_NDM_, bla_VIM_, and bla_OXA-48_ were relatively rare and mainly found in clades B2 and B3 than in A and B1. Comparatively, other Enterobacteriaceae species (*E. coli, K. pneumoniae*, and *S. marcescens*) had strains with richer resistomes diversity than *C. freundii* clades A and B1, including chromosomal mutations and MDR efflux pumps. However, Clades B2 and B3 had comparable resistome diversity and abundance to the above-mentioned Enterobacteriaceae species, in which CMY was well-nigh absent except in *E. coli* (Fig. 1).

A local outbreak of *C. braakii* was observed in the UK (in humans), with closely related strains being isolated from biosolids (Canada), carrots (Germany), chicken (China) and beef (Canada) (Fig. S1A-B). *C. amalonaticus* was also found in humans, animals, plants and the environment (Fig. 2): *bla*_CMY_ was not common in this species, but *bla*_SED_ and *OqxA/B* were almost conserved in clades B1 and C, with clade B2 (all from France) having the richest resistome repertoire that included *bla*_TEM_, *bla*_SHV_, *bla*_NDM_ and *mcr*-9.1 (Fig. 2). *C. koseri* had a relatively limited resistome diversity, with *bla*_MAL-1/2_ (clade B2), *bla*_CKO_ (clade B3) and *fos*A7 (clade B2) being the commonest ARGs. Comparatively, the other Enterobacteriaceae species (e.g. *E. coli, K. pneumoniae, Enterobacter spp., Serratia spp*., and *Providencia spp.)* had richer resistome diversity than *C. koseri* (Fig. 3).

**Figure 2.**
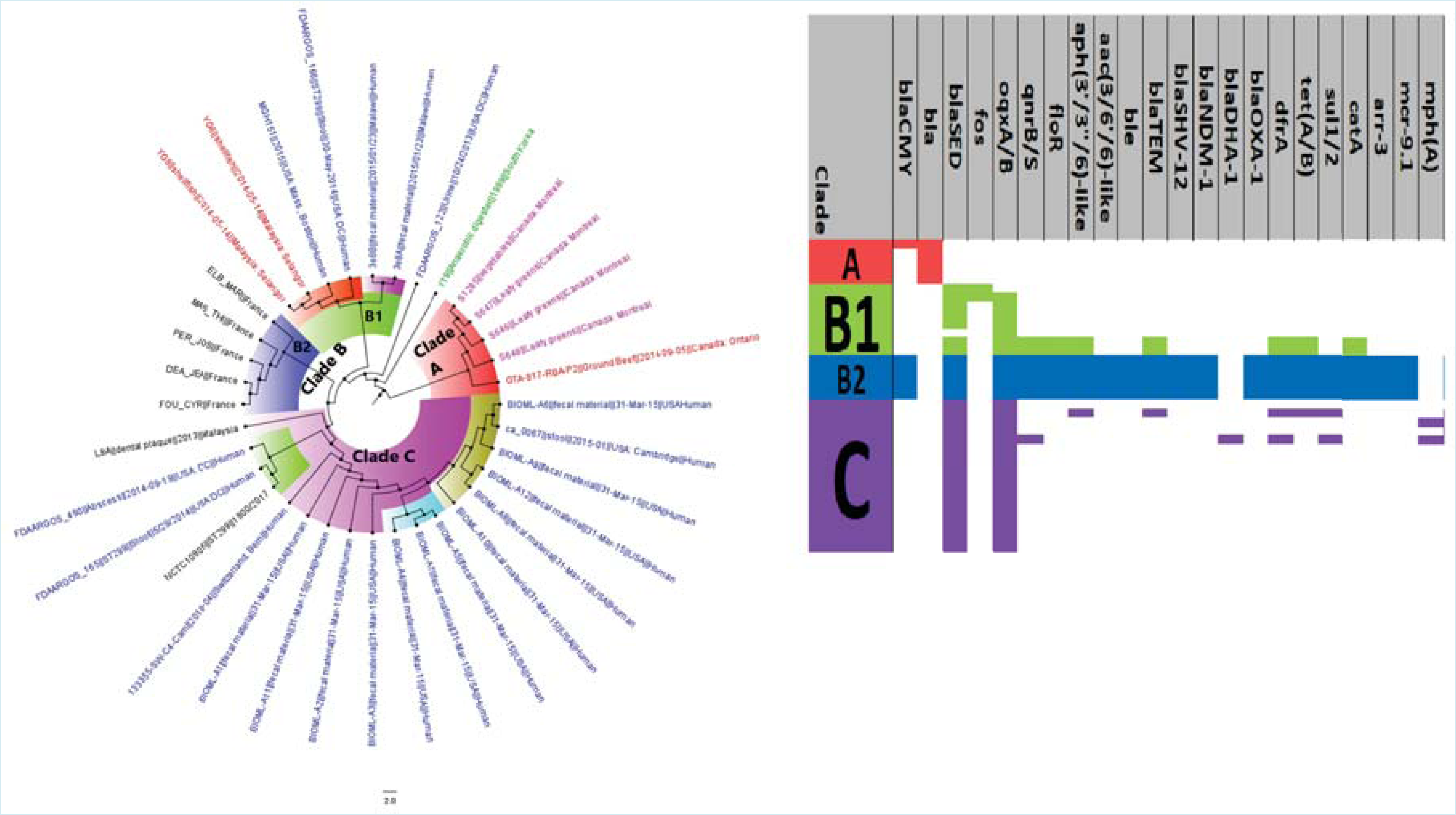
Evolutionary epidemiology and resistome of global *Citrobacter amalonaticus* isolates. *C. amalonaticus* strains clustered into clades A (red highlight), B1 (green highlight), B2 (blue highlight) and C (mauve highlight); clade B2 had very rich resistome repertoire and were all from France, but the other clades had very few resistance genes. Strains from humans (blue labels), animals (red labels), plants (purple/mauve labels) and the environment (green labels) were found in the same clade/cluster. *Bla*_SED_ and *oqxAB* were almost conserved in these genomes.

**Figure 3.**
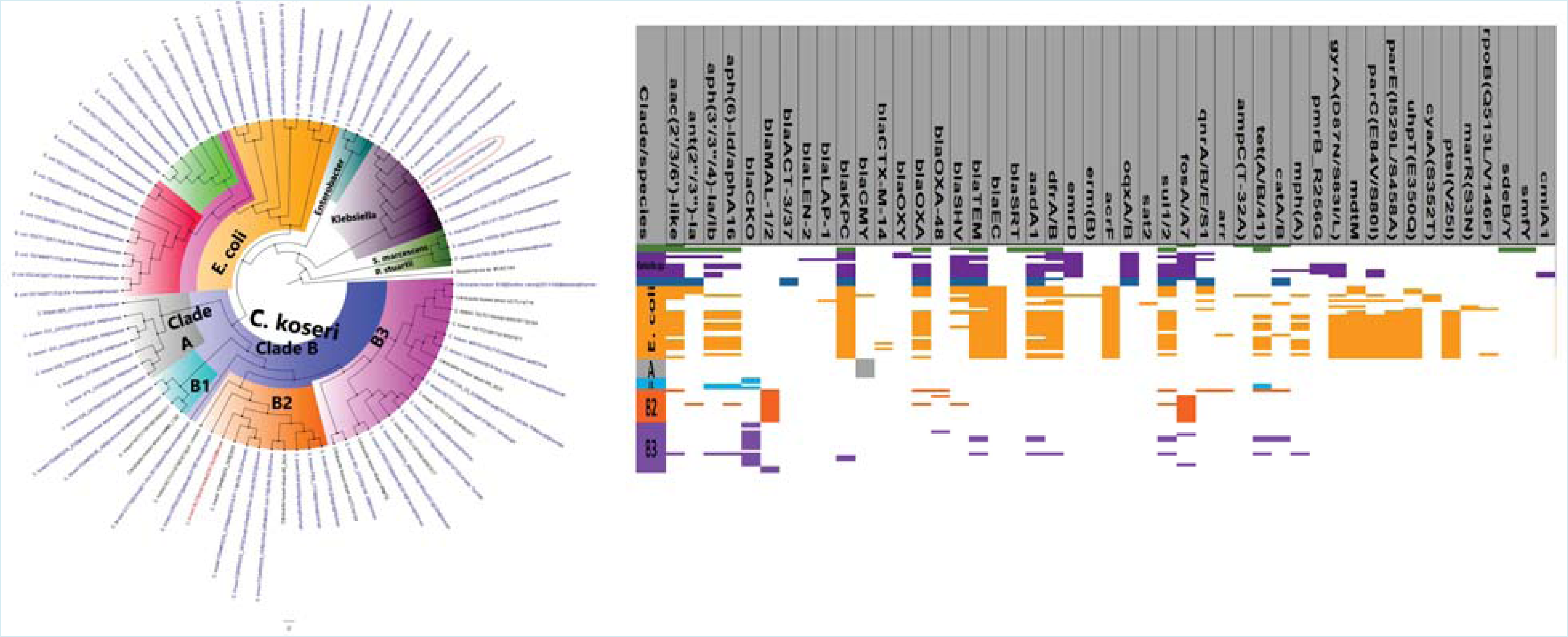
Evolutionary epidemiology and resistome of global *Citrobacter koseri* isolates. *C. koseri* strains clustered into clades A (grey highlight), B1 (light blue highlight), B2 (orange highlight) and B3 (mauve highlight). Strains from humans (blue labels) and animals (red labels) were found in the same clade/cluster. *Bla*_CKO_ and *bla*_MAL_ were almost conserved in these genomes.

#### Enterobacter species

The resistomes of *E. steigerwaltii* and *E. oharae* were extraordinarily rich and diverse, although clade A of *E. steigerwaltii* had lesser resistome abundance and diversity compared to clades B and C (Fig. 4). *E. oharae* only clustered in clade B of *E. steigerwaltii* and had most *bla*_CTX-M_, *bla*_SHV_, *bla*_NDM_, *bla*_OXA-48_ and *bla*_VIM_ ARGs than *E. steigerwaltii* clade C. *bla*_ACT_ was present in almost all *Enterobacter spp*. strains whilst other AmpCs such as *bla*_LAP_, *bla*_SCO-1_, *bla*_SFO_, *bla*_TMB_, *bla*_DHA_, and *bla*_CARB_ were virtually absent. *E. cloacae* strains clustered closely with *E. steigerwaltii* clades B and C. overall, *E. steigerwaltii* and *E. oharae* strains were richly endowed with clinically important ARGs including carbapenemases, ESBLs, *aac(6’)-like, aac(3’)-like, aph(3’/3”)-like, aadA, dfrA, catA, fosA, oqxAB, qnrA/B/S/D, sul1/2*, and tet(A/B/D). *Mcr-9* genes were particularly abundant in clade B than in clades A and C of *E. steigerwaltii/cloacae* (Fig. 4).

**Figure 4.**
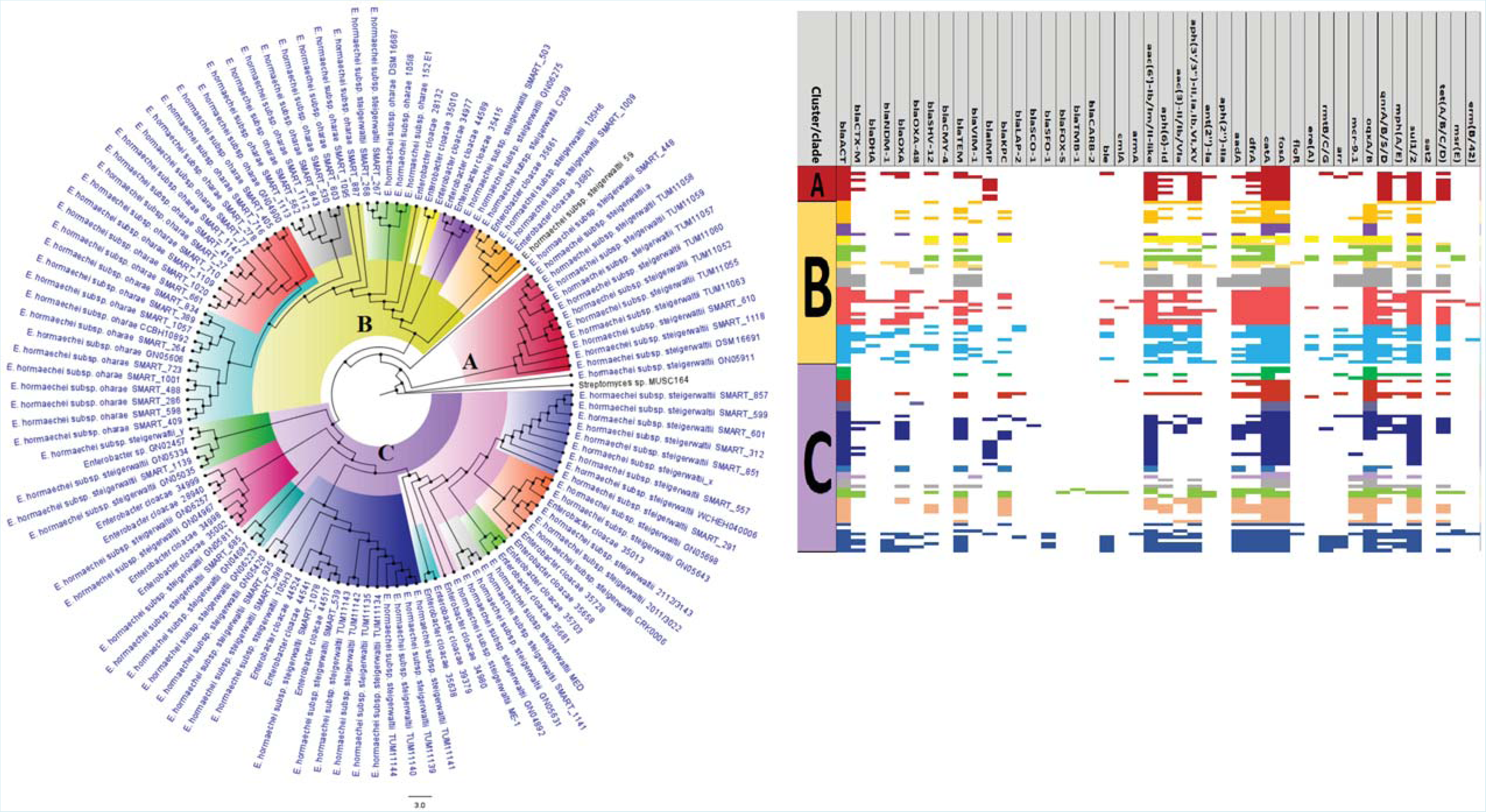
Evolutionary epidemiology and resistome of global *Enterobacter steigerwaltii/oharae* isolates. *E. steigerwaltii/oharae* isolates clustered into clades A, B and C, which were all obtained from humans from countries distributed across the globe. Clades B and C had very rich resistome repertoire; *bla*_ACT_ was conserved in these genomes..

*E. cloacae* strains clustered with all *E. xiangfangensis* strains except clade A1, from which it was evolutionarily distant (Fig. 5). ARGs such as *bla* _ACT_, *catA/B, OqxA/B* and *fosA* were universally present in almost all the *E. xiangfangensis* and *E. cloacae* strains. Furthermore, *E. xiangfangensis* clade A1 had the least ARGs diversity and abundance whilst *E. cloacae* A1 and the remaining *E. xiangfangensis* (clades A2 and B) were richly endowed with multiple ARGs such as *bla*_KPC_, *mcr-9, bla*_CTX-M_, *bla*_OXA_, *bla*_NDM_, *bla*_TEM_, and genes mediating resistance to fluoroquinolones (aac(6’/3’)-like, *qnr*A/B/S etc.) and aminoglycosides (*armA, rmtC/G, aadA, aph*(4)-, *aph*(3’/3”)- and *aph*-(6’)-like) as well as *tet* (A/B/D), *dfrA*, and *arr* (particularly in clades A2 and B). Thus, the resistomes diversity and abundance of *E. xiangfangensis* and *E. cloacae* were comparable to that of *E. oharae* and *E. steigerwaltii* (Fig. 4-5).

**Figure 5.**
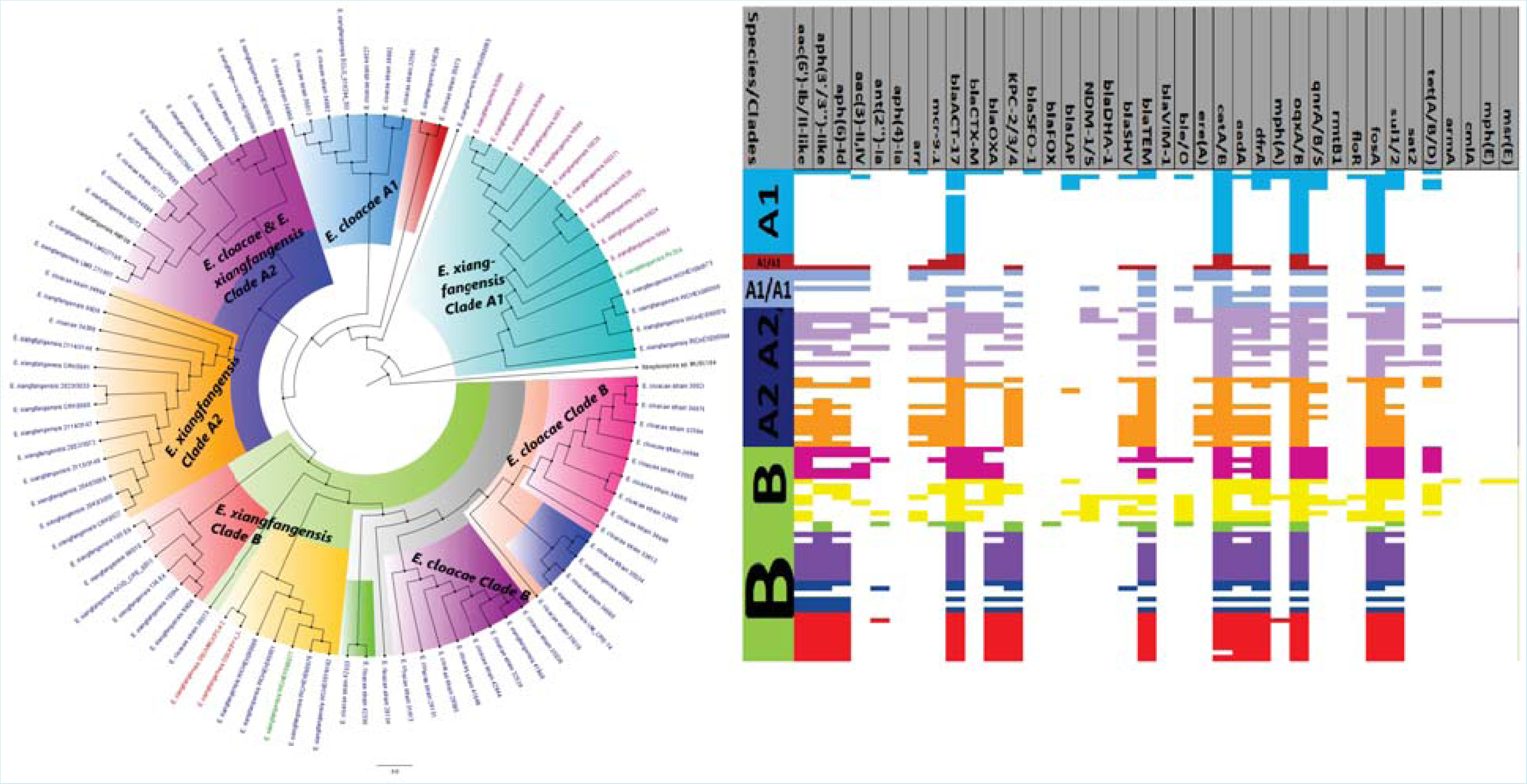
Evolutionary epidemiology and resistome of global *Enterobacter xiangfangensis* isolates. The *E. xiangfangensis* contained *E. cloacae* genomes and they clustered into clades A1, A2 and B, with clades A2 and B having rich and diverse resistome repertoire; these clades were distributed globally from humans (blue labels) and animals (red labels). *bla*_ACT_ was conserved in these genomes.

An *E. hormaechei* outbreak was observed in Germany in 2017, evolving with the spread (clade B) (Fig. 6 & S2). *E. steigerwaltii* and *oharae* were mainly isolated from humans whilst *E. xiangfangensis* and *E. hormaechei* were from humans, plants (*E. xiangfangensis* from rice in India), animals and the environment. Strains of closely related (i.e. close evolutionary distance) *E. hormaechei* in clade B were from the US, South Africa, Colombia, China, Germany, Australia and Lebanon from humans, animals, and plants (Fig. 4-6). Other closely related *E. hormaechei* strains from humans, animals, plants, and the environment were found in different countries, showing a gradual evolution of strains emanating from a common ancestor and spreading across countries through different hosts. These observations were made in *E. hormaechei* clades B and C, and represented local and international outbreaks spanning UK, USA, China, Serbia, Japan etc. (Fig. 6 & S2).

**Figure 6.**
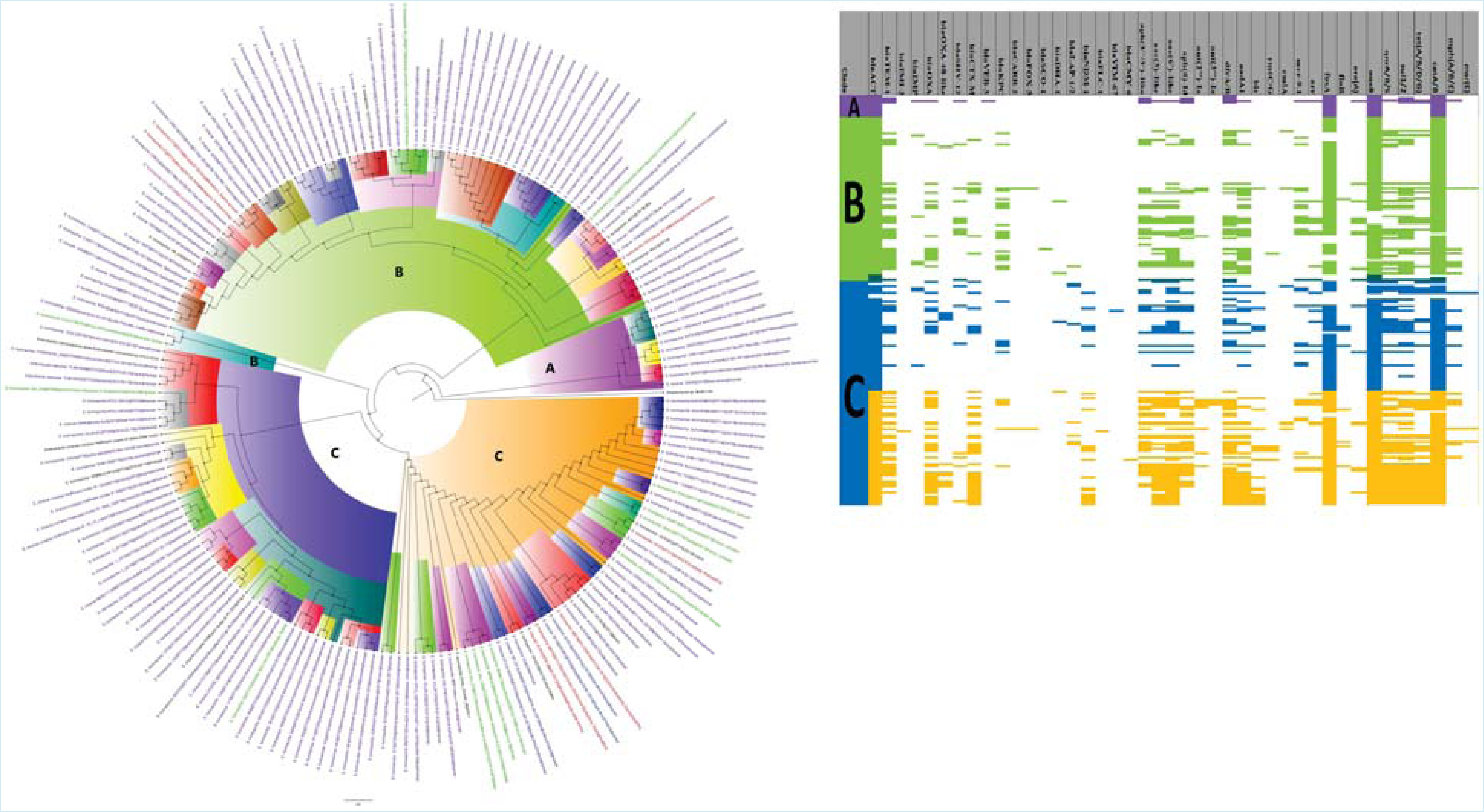
Evolutionary epidemiology and resistome of global *Enterobacter hormaechei* isolates. The *E. hormaechei* isolates clustered into three main clades A, B and C (with distinct highlights) that contained strains distributed globally from humans (blue labels), and animals (red labels), plants (purple/mauve labels) and the environment (green labels). Clades B and C contained diverse and rich resistome repertoire. *bla*_ACT_ was conserved in these genomes.

Moreover, *bla*_ACT_, *fosA*, OqxA/B and *cat*A/B were almost conserved in almost all the clades of *E. hormaechei*. As observed with the other *E. hormaechei* subsp. (*oharae, steigerwaltii* and *xiangfangensis*), *E. hormaechei* clades B and C were richly endowed with ARGs than some clade A strains (except clade A in Fig. S2A). Specifically, clades B and C strains were relatively enriched with *bla*_TEM-1_, *bla*_OXA_, *bla*_CTX-M_, *aph(3’/3”)-like, aac(3’)-like, aac(6’)-like, dfrA/B, mcr-9, arr, qnrA/B/S, sul-1/2*, and *tet*(A/B/C/D). Further, clade B was mostly rich in *bla*_KPC_ and *bla*_NDM_ whilst clade C was rich in *bla*_NDM_ and *ble*. A substantial number of clade A strains also had *mcr-9* and *bla*_TEM_ (Fig. 6 & S2).

*E. oharae* was evolutionarily closer to *E. xiangfangensis*, clustering on the same branches, with a relatively few *E. oharae* strains clustering with *E. steigerwaltii*. However, *E. steigerwaltii* was mostly distant from *E. oharae* and *E. xiangfangensis*, with a few *E. xiangfangensis* strains (from rice in India) clustering within *E. steigerwaltii* clades A and C (Fig. 2SB). Figure S2B summarises the ARGs in *E. hormaechei* and its subspecies: *bla*_ACT_, *fosA, OqxAB, qnrA/B/D/S*, and *catA/B* were conserved whilst *bla*_OXA_, ble, *bla*_SHV_, *bla*_TEM_, bla_KPC_, *bla*_NDM_, *bla*_VIM_, *mcr, tet, mph(A), sul-1/2, dfrA*, and fluoroquinolone and aminoglycoside ARGs were richly abundant, particularly in clusters IV, V and VII.

#### K. variicola

*K. variicola* strains were isolated mainly from humans, with some being from animals, plants, and the environment. On the individual branches/clades were closely related strains with very close evolutionary distance but disseminated across countries, suggesting international dissemination of that clade and showing little evolution during the spread from host to host, e.g., clades A3, A4, A5, A6, B1, B2 and C (Fig. 7 & S3). As well, local outbreaks of closely related strains in the USA, Bangladesh, Canada, and Germany were observed. A reassessment of the *K. variicola* tree with genomes of *K. pneumoniae* and *K. quasipneumoniae* largely confirmed the initial clustering of the *K. variicola* strains, with only a few rearrangements of some strains within different clades (Fig. S3A and S3B).

**Figure 7.**
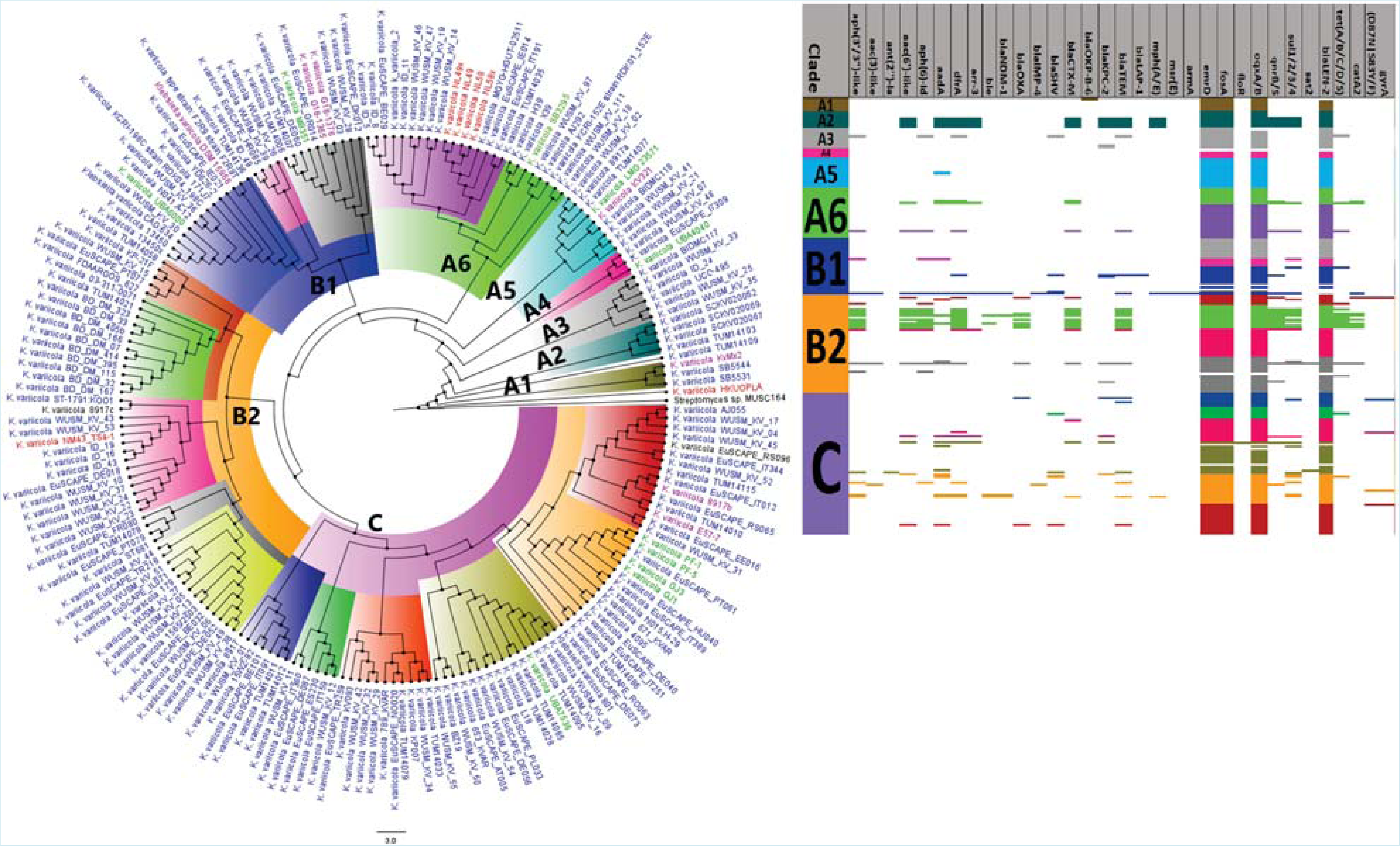
Evolutionary epidemiology and resistome of global *Klebsiella variicola* isolates. The *K. variicola* strains clustered into nine clades viz., A1, A2, A3, A4, A5, A6, B1, B2 and C, which were highlighted with distinct colours and were isolated from countries around the globe. The clades contained strains distributed globally from humans (blue labels), animals (red labels), plants (purple/mauve labels) and the environment (green labels). Besides a few strains in clade B2, the other strains contained very few resistance genes. *bla*_LEN_ was conserved in these genomes.

Notably, *K. variicola* had fewer resistome diversity and abundance than *Citrobacter spp., Enterobacter spp*., and *K. pneumoniae spp*. Conserved within the *K. variicola* genomes were *emrD, fosA, OqxAB* and *bla*_LEN-2_, whilst the other ARGs were sparse. Whilst *bla*_SHV_ was conserved in *K. pneumoniae*, it was virtually absent in *K. variicola;* further, *bla*_LEN_ was present in the latter but absent in the former. *K. variicola* clade B2 strains (on branch VIII), specifically those from Bangladesh, had richer and more diverse resistomes than the other *K. variicola* clades/clusters. Whilst *mcr* and *bla*_NDM_ were virtually absent in these genomes, *bla*_KPC_ occurred in substantial abundance. These suggest that *K. variicola* is least likely to be MDR and/or harbour ESBLs, carbapenemases, *mcr* and other clinically important ARGs compared to other Enterobacteriaceae species (Fig. 7 & S3).

#### M. morganii

There were three*M. morganii* clusters/clades viz., A, B and C, which were mainly from humans with a few in clades A and C being from animals (Fig. 8). Within each clade are closely related strains with very close evolutionary distance that were distributed across several countries; indeed, strains of the same clone were found in different countries, suggesting international dissemination of the same clones. The branching order of the trees within each clade shows the gradual evolution of the strains as they moved from host to host. *bla*_DHA_ and *catA/B* genes were almost conserved in almost all the *M. morganii* genomes. Other highly abundant ARGs in the *M. morganii* strains were *sul-1/-2, tet*(A/B/D/Y), *bla*_OXA-1_, *ble, dfrA1, aadA, aac(3)-like* and *aph(3’/3”)-like*. Notably, clade B had more ARG diversity and abundance than clade C and A; clade A had the least diversity and abundance of ARGs (Fig. 8).

**Figure 8.**
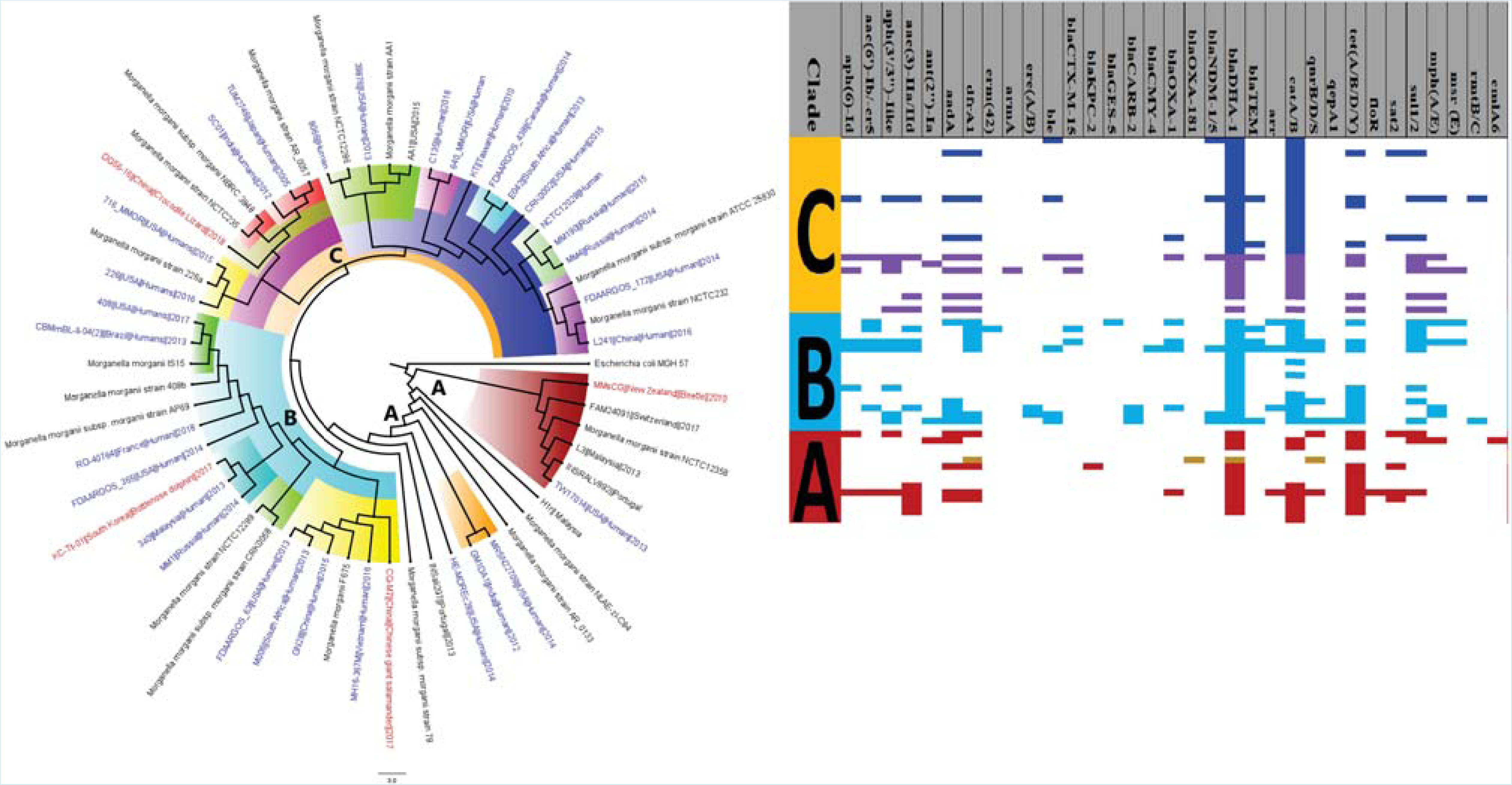
Evolutionary epidemiology and resistome of global *Morganella morganii* isolates. The *M. morganii* strains clustered into three clades, A (red highlight), B (light blue highlight) and C (yellow/gold highlight), containing isolates obtained globally from humans (blue labels) and animals (red labels).

#### P. mirabilis

*P. mirabilis* clustered into three major clades and included isolates from humans, animals, and the environment. Within the three clades were sub-clades consisting of closely related strains from the same as well as different countries, showing local and international outbreaks involving human and animal hosts, and in some cases environmental mediators (Fig. 9). Specifically, local outbreaks were seen in the USA (clades A3, B2 and B3), France (clade C2) and Japan (C3) whilst international dissemination of clades A2, A3, A4, B, C2 and C3 were observed. Clade C2 had the richest abundance of resistomes, with all the members having a uniform/conserved diversity of the same ARGs, except for *bla*_CTX-M_, *bla*_OXA_, *bla*_TEM_, and *Inu*(F/G). Clade C3 had the 2^nd^ most abundant but more diverse ARGs than C2. Notably, clade B and its subclades had lesser ARGs than clades A and C. *CatA* and *tet*(A/B/D/Y) ARGs were virtually conserved in all the clades whilst *aadA, aac(3’)-like, aph(3’/3”)-like, aph(6’)-like, dfrA7, sat2*, and *sul-1/2/3* were substantially prevalent in all the clades. In particular, bla_CARB2_ and *florR* were highly conserved in clade C2; *florR* was however less abundant than *bla*_CARB_. Notably, *mcr* was almost absent except in a few strains in B3. As well, *bla*_CTX-M_, *bla*_CMY_, *bla*_NDM_, *bla*_OXA-1_, *bla*_TEM_ and *ble-O/Sh* were mainly found in clades A and C3, with traces in B and C1. Thus, ESBLs, carbapenemases and *mcr* genes were relatively less common in *P. mirabilis* strains, compared to other Enterobacteiaceae (Fig. 9).

**Figure 9.**
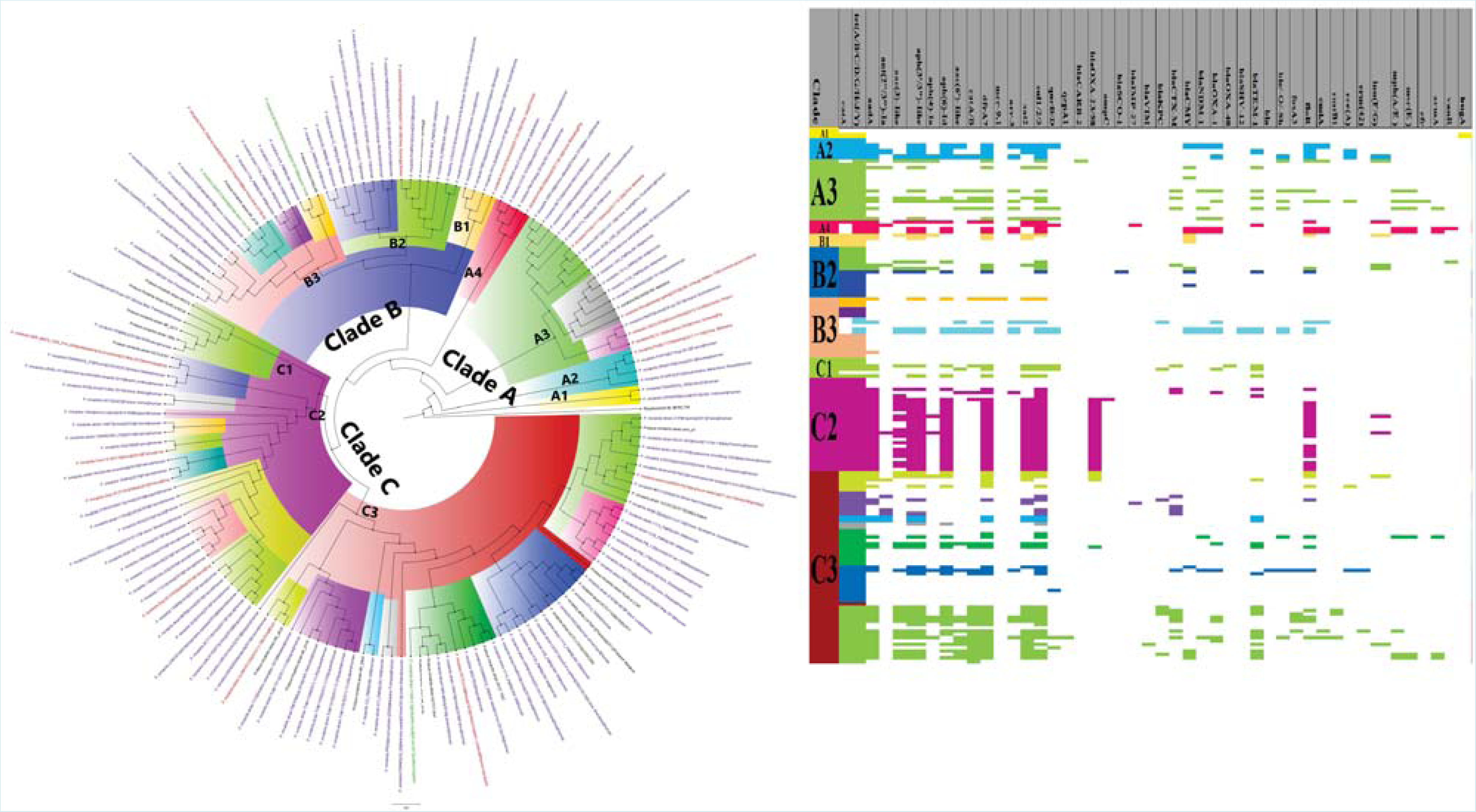
Evolutionary epidemiology and resistome of global *Proteus mirabilis* isolates. The *P. mirabilis* isolates clustered into 10 clades, A-A3, B1-B3, and C1-C3 (shown with different highlights), which contained diverse and abundant resistomes with conserved *catA* and *tet* genes. The clades contained strains distributed globally from humans (blue labels), animals (red labels), plants (purple/mauve labels) and the environment (green labels).

#### Providencia species

There were five *Providencia spp*. viz., *stuartii, rustigianii, alcalifaciens, heimbachae*, and *rettgeri*, with *P. rettgeri* being most isolated and branching into two clades, A and B (Fig. 10). The other species had single clades. *Providencia spp*. were isolated from animals, humans, and the environment. Highly similar strains of the same species were found across countries (*P. stuartii, P. alcalifaciens, P. heimbache* and *P. rettgeri*) and within countries (*P. rettgeri* clade A). The distinction between the various species of Providencia was depicted by the clustering patterns on the tree as strains of the same species clustered together; *Providencia spp*. was closest evolutionarily to *S. marcescens* whilst *E. coli* was closest to *Citrobacter spp*.,and *E. hormaechei*. As well, *K. pneumoniae, K. aerogenes*, and *K. michiganensis* clustered together with relatively short evolutionary distance (Fig. 10).

**Figure 10.**
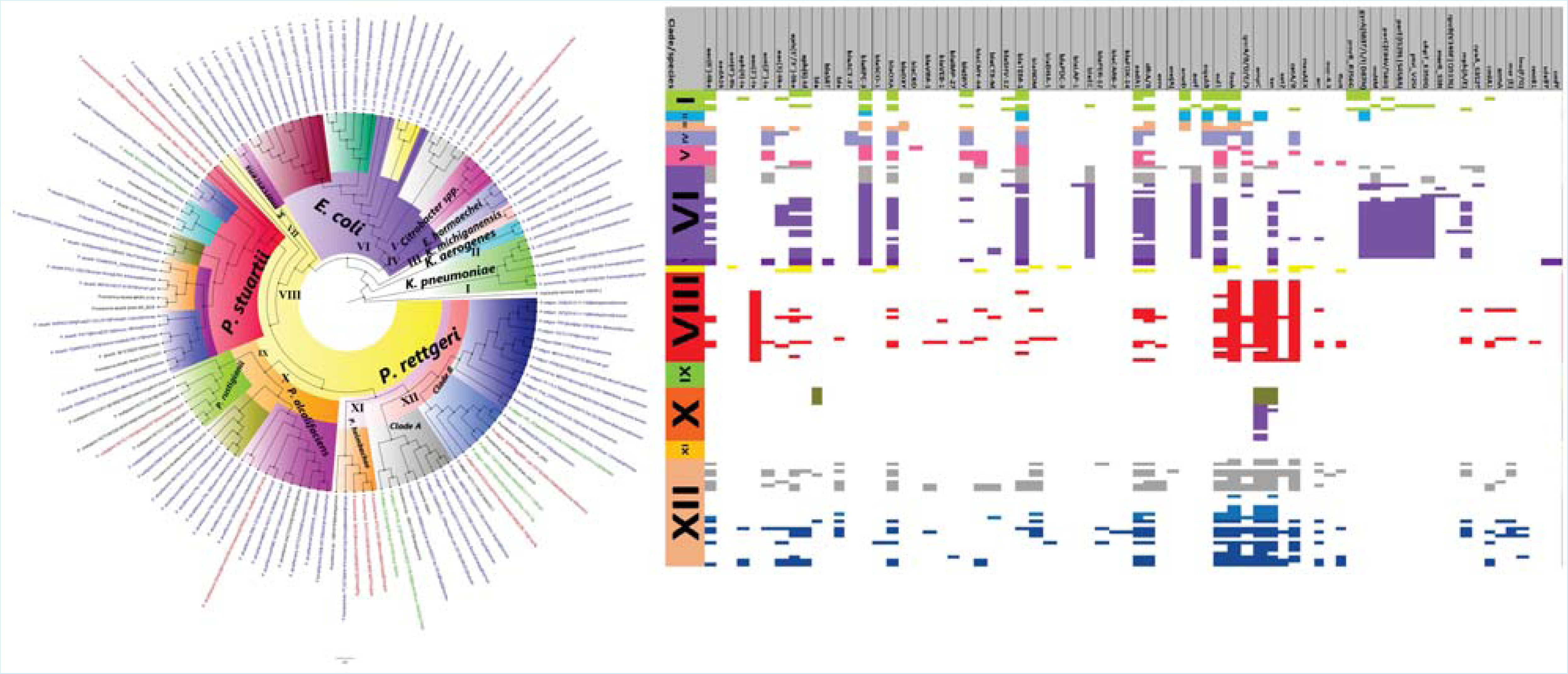
Evolutionary epidemiology and resistome of global *Providencia spp*. isolates. The *Providencia spp*. clustered into 12 branches, with *P. stuartii, P. rustigianii, P. alcalifaciens, P. heimbachei* and *P. rettgeri* clustering into branch VIII to XII respectively. *P. rettgeri* clustered into clades A and B, consisting of globally distributed isolates. *P. stuartii* and *P. rettgeri* contained richer and more abundant resistomes than the other *Providencia* strains and contained strains distributed globally from humans (blue labels), animals (red labels), plants (purple/mauve labels) and the environment (green labels).

The other Enterobacteriaceae species had more ARGs than *Providencia spp*. Within *Providencia spp., P. stuartii* and *P. rettgeri* were most endowed with richer and more diverse resistomes whilst *P. alcalifaciens* and *P. rustigianii* were almost bereft of ARGs. Common ARGs within *P. stuartii*, were *aac(2’)-Ia*, aph(3)-like, *bla*_OXA_, *bla*_TEM_, *aadA1, dfrA/B, sul1, fosA, ampC, tet* and *catA/B* whilst *P. rettgeri* had *aac(6’)-like, aph(3’/3”)-like, bla*_SRT_, *bla*_OXA_, *bla*_NDM_, *aadA, dfrA/B, OqxAB, sul1, qnrA/B/D/E/S, ampC, catA/B, tet, arr*, mph(A/E), *cmlA*, msr(E), and Inu(F/G). Hence, *P. rettgeri* has the most abundant and diverse ARGs than all other *Providencia spp*. (Fig. 10).

#### Phylogeography

North America (particularly USA) and Europe (particularly Western and Southern Europe) had the highest concentration of the various species, followed by South East Asia, South America (particularly Brazil and Colombia) and South Africa. There were sparse reports on these species from Australasia, the Middle East and Africa (except South Africa) and the Caribbean (Fig. 11).

**Figure 11.**
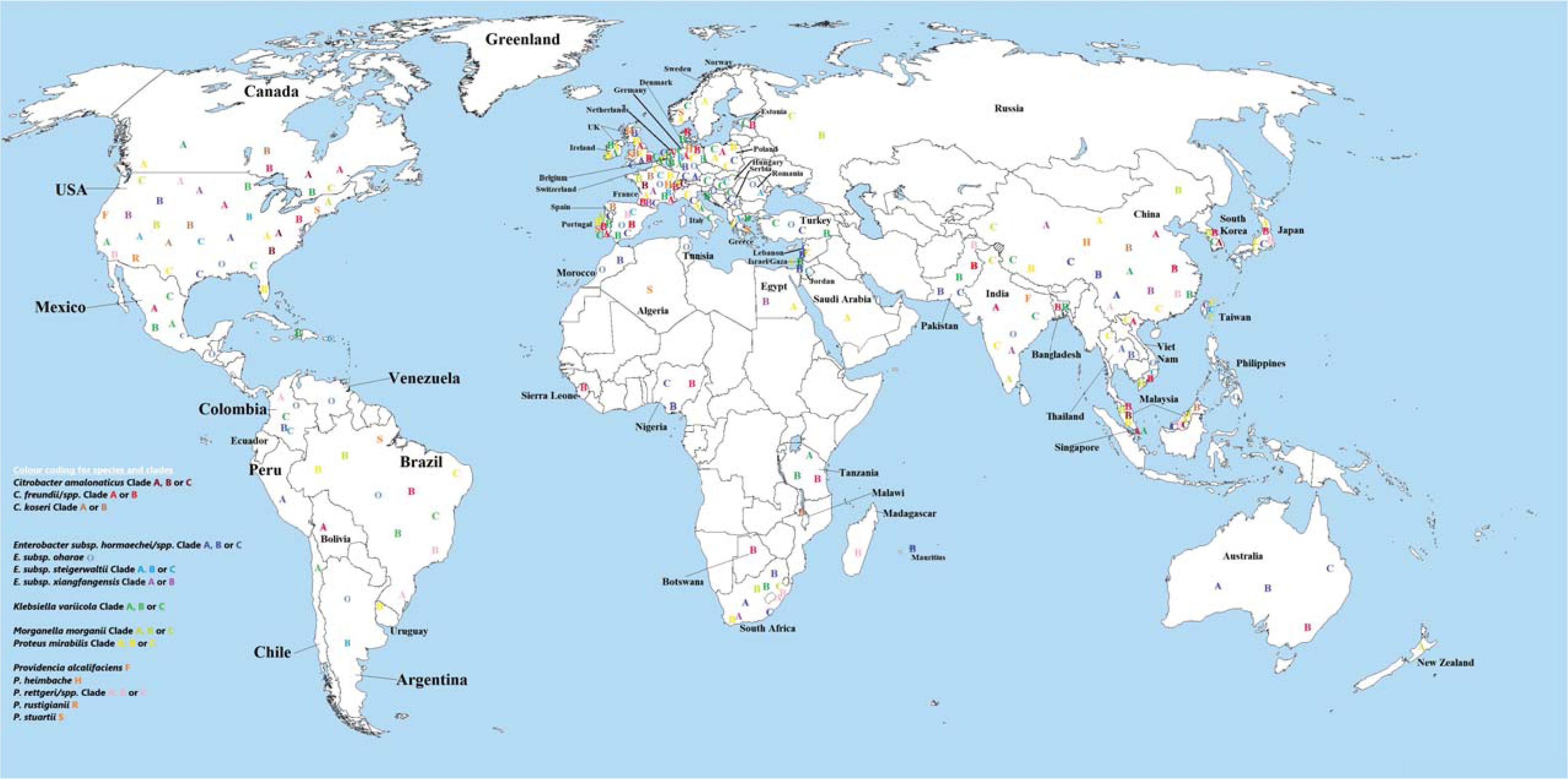
Global phylogeography of *Citrobacter freundii, Citrobacter amalonaticus, Citrobacter koseri, Enterobacter hormaechei subsp. hormaechei, xiangfangensis, steigerwaltii* and *oharae, Klbesiella variicola, Morganella morganii, Proteus mirabilis*, and *Providencia spp*. Most of these genomes were obtained from USA, Europe, South-East Asia and South America in a descending order of frequency. *C. freundii, Enterobacter spp., K. variicola*, and *P. mirabilis* had more diverse distribution across the globe. Each species is designated with a different colour code.

*C. freundii* clade A was distributed mainly in North America, Europe, and South-East Asia whilst clade B was found worldwide on almost all continents. *C. amalonaticus* clade A was found in North America and South Korea; clade B was found in Malawi, USA, Malaysia, and France whilst clade C was only found in Malaysia, USA and Switzerland. *C. koseri* clade A (USA) and clade B (USA, Spain, UK, France, Canada, China and Malaysia) were relatively less reported with clade B being more widely distributed globally (Fig. 11).

*E. oharae* strains and *E. hormaechei* clades A, B and C were globally disseminated, with clade C being most widely distributed, followed by clades B and A. *E. steigerwaltii* clade C was more globally distributed than clades A (USA, Japan and Europe) and B (Argentina, USA, France and Germany). Although *E. xiangfangensis* was globally disseminated, the respective clades A (China, India, USA, S. Africa, and France) and B (France, USA, Egypt, and China) were reported from very few countries (Fig. 11).

*K. variicola* strains, particularly clades B and C, were of wide geographical distribution; clade A was found in relatively fewer countries globally. *M. morganii* strains were found in North America, Europe (including Russia), and South-East Asia, with clades B and C being found in South Africa. *P. mirabilis* clades A, B and C were found globally, with clade C being reported in most countries. *P. rettgeri* strains had the widest global distribution and reports from most countries among the *Providencia spp*., followed by *P. stuartii. P. rettgeri* clade A (China, Brazil, South Africa, Colombia, and USA) was found in fewer countries than clade B. *P. stuartii* strains were of global distribution whilst *P. rustigianii* (UK and USA), *P. alcalifaciens* (USA and India) and *P. heimbachae* (France, China, and Germany) were not (Fig. 11).

### Frequency distribution of ARGs per species

#### MCR ARGs

*Mcr-9.1* ARGs were the commonest *mcr* variants identified, with very few *mcr-1* and *mcr-3* being found in *E. hormaechei* and *C. freundii (mcr-1, -3* and *-10)* and a single *mcr-4.3* gene being found in *P. rettgeri*. Notably, the highest prevalence of *mcr-9* was in *E. hormaechei* (n=67 *mcr-9* genes), *E. steigerwaltii/oharae* (n=32 *mcr-9* genes) and *E. xiangfangensis/cloacae* (n=19 *mcr-9* genes), followed by *C. freundii* (n=19), *C. amalonaticus* (n=5) and other *Citrobacter spp*., some of which had very few or no *mcr* genes (Fig. S4-S9).

#### Carbapenemases

One of the most prevalent carbapenemase among the species was KPC, with KPC-2 (n=123), KPC-3 (n=97), KPC-4 (n=14), and KPC-6 (n=1) being common. KPC-2 was higher in all the species except in *E. xiangfangensis* for which KPC-3 was more abundant (n=35) than KPC-2 (n=9). KPC was most prevalent in *E. hormaechei/oharae/steigerwaltii/xiangfangensis* and *C. freundii* strains whilst *C. amalonaticus, C. koseri*, and *Providencia spp. had no* KPC ARGs. After Ambler class A KPC, Ambler class D OXA-48-like serine carbapenemases (n=65) were also very prominent in all species except *C. amalonaticus, E. xiangfangensis* and *K. variicola*, with other 0XA-48 variants such as OXA-181 (n=1, *M. morganii)* and OXA-396 (n=1, *P. rettgeri)* being relatively scarce. *C. freundii* (n=43), *E. hormaechei* (n=10), *E. oharae/steigerwaltii* (n=9) and *C. koseri* (n=3) had OXA-48 genes whilst OXA-58 (n=2) and OXA-23 (n=23) were only found in *P. mirabilis*. Other class A serine carbapenemases i.e., GES-5 (n=1, *M. morganii)* and IMI-2 (n=1, *E. hormaechei)*, were also rare (Fig. S4-S9).

NDM was the commonest class B carbapenemase (n=159), followed by IMP (n=97) and VIM (n=83). NDM-1 (n=137) was the most prevalent variant and was found in *E. hormaechei* (n=68), *C. freundii* (n=15), *E. steigerwaltii* (n=13), *Providencia spp*. (n=12), *P. mirabilis* (n=10), *E. xiangfangensis* (n=6), *C. amalonaticus* (n=5), *M. morganii* (n=5), and *K. variicola* (n=3), with NDM-5 (n=16; 12 in *E. hormaechei*, 3 in *E. xiangfangensis*, and 1 in *P. mirabilis*), NDM-7 (n=3 in *E. hormaechei*) and NDM-9 (n=3 in *K. variicola*) being less prevalent. IMP-1 (n=71; 67 in *E. hormaechei* and 4 in *C. freundii)*, IMP-8 (n=14; 13 in *C. freundii*, 1 in *E. steigerwaltii)*, IMP-4 (n=9; 5 in *C. freundii*, 4 in *E. hormaechei)*, IMP-27 (n=2 in *P. mirabilis)* and IMP-13 (n=1 in *E. oharae)* were the identified variants. VIM-1 (n=65; 26 in *C. freundii*, 14 in *E. steigerwaltii*, 21 in *E. hormaechei*, 1 in *E. xiangfangensis*, 2 in *Providencia spp*., 1 in *P. mirabilis)*, VIM-4 (n=12; 5 in in *E. steigerwaltii*, 5 in *E. hormaechei*, 2 in *C. freundii)*, VIM-2 (n=2 in *Providencia spp*.), VIM-31 (n=2; *E. steigerwaltii* and *E. hormaechei)*, VIM-5 (n=1 in *E. hormaechei)*, and VIM-67 (n=1 in *E. hormaechei*) were identified in the strains (Fig. S4-S9).

#### ESBLs and ampCs

TEM was the commonest ESBL and TEM-1 was the most common variant to be identified in all species; particularly, TEM-1 was most abundant in *Enterobacter spp*. and *C. freundii*. OXA (particularly OXA-1, -9, and -10), CTX-M (particularly CTX-M-15) and SHV (particularly SHV-12) were also common in almost all species in a descending order of prevalence, but were very abundant in *Enterobacter spp*., specifically *E. hormaechei; C. koseri, C. amalonaticus, P. mirabilis*, and *M. morganii* had relatively low abundance of SHV, OXA and CTX-M genes. Other ESBLs genes such as *bla*_SCO_, *bla*_LAP_, *bla*_VEB_, *bla*_TMB_, *bla*_SFO_, *bla*_SMB_, *bla*_CARB_, *bla* etc. were also rare in the various species (Fig. S4-S9).

AmpC ARGs were basically strain-specific, with ACT, LEN, CMY, DHA, CKO/MAL and SED being conserved in *Enterobacter spp., K. variicola, C. fruendii, M. morganii, C.koseri* and *C. amalonaticus* (except clade A) respectively; FOX was rare (Fig. S4-S9).

#### Aminoglycoside ARGs

ARGs mediating resistance to aminoglycosides such as *aadA, aph(2”)-like, aph(3’)-like, aph(4’)-like, aac(6’)-like, aac(3)-like, aph(6)-like*, and *ant(2”)-like* were abundantly prevalent in almost all the clades of *C. freundii, Enterobacter spp*., and *P. mirabilis* and sparsely abundant in the other species; *aadA, aph(4’)-like, aac(6’)-like* and *aac(3’)-like* ARGs were most common. 16S rRNA Methyltransferases such as *rmtB1 (E. steigerwaltii* and *P. mirabilis*,), *rmtC/G (E. hormaechei, C. freundii* and *M. morganii)*, and *armA (C. freundii, E. xiangfangensis, P. mirabilis, P. stuartii, M. morganii* and *P. rettgeri*) were rare (Fig. S4- S9).

#### Fluoroquinolone ARGs

*Aac(6’)-like, OqxAB, QnrA/B/D/S* and *qepA* ARGs were identified, albeit *qepA (M. morganii)* was rare and *Oqx*A was less prevalent than *Oqx*B in all but one species. Notably, *OqxAB* were virtually absent in *C. fruendii* clades. whilst chromosomal mutations in *gyr*AB and *parCE* were only observed in *K. variicola* (Fig. S4-S9).

#### Other ARGs

Chloramphenicol ARGs, *cmlA, catA* and *catB* were found in all the species, albeit *cmlA* was relatively rare in all the species whilst *catA/B* were conserved in *Enterobacter spp*. and *M. morganii; catA* was more prevalent than *catB. catA/B* were also abundant in *Citrobacter spp., P. mirabilis* and *Providencia spp*. (Fig. S4-S9). *Sulphamethoxazole-trimethoprim* ARGs, *sul-1/2/3* and *dfrA*, were enriched in *Providencia spp., P. mirabilis, M. morganii, Enterobacter spp., C. koseri (dfrA was virtually absent), C. amalonaticus* clade B2, and *C. freundii. Sul1* was more prevalent than *Sul2*, with *Sul3* being relatively rare whilst the *dfrA* variants were remarkably diverse. Indeed, both *sul1, sul2* and/or *sul3* as well as several *dfrA* variants were present concurrently in some single strains (Fig. S4-S9; Tables S2).

Several tetracycline ARGs such as *tet*(A), *tet*(B), *tet*(C), *tet*(D), *tet*(G), *tet*(*J*), *tet*(S), *tet*(Y), and *tet*(41), were present in all the species. Notably, *tet*(A), *tet*(B), and *tet*(D), were highly enriched in the various genomes with a descending order of frequency; *tet*(J) was most prevalent in *P. mirabilis* (Fig. S4-S9).*fosA* variants were present in all the species except *M. morganii*, but were most enriched and conserved in *Enterobacter spp. K. variicola, P. stuartii* and *P. rettgeri*. As well, *ere*(A), *emr*(D), *erm*(B), *msr*(E), *mph*(A) and *mph*(E) macrolide ARGs were common in the various species, with *mph*(A) being richly abundant; *mph*(E) and *msr*(E) were enriched in *C. freundii, Providencia spp*. and *P. mirabilis, emr*(D) was abundant in *K. variicola* whilst *ere*(A) was abundantly enriched in *E. hormaechei* and *E. xiangfangensis*. Rifamycin ARG, *arr*, was identified in the various species, represented by *arr-S arr-2* and *arr-1* in all the species (Fig. S4-S9).

## Discussion

Among Gram-negative bacterial species, *Pseudomonas aeruginosa, Acinetobacter baumannii*, and Enterobacteriaceae such as *K. pneumoniae, E. coli, S. enterica, Vibrio cholerae*, and *Shigella spp*. are commonly isolated and implicated in nosocomial (anthroponotic), zoonotic and water/food-borne infections with MDR, extensively and pandrug resistant (XDR and PDR respectively) phenomes ^4,26,28,37,38^. However, other less isolated Enterobacteriaceae species such as *Citrobacter spp., Enterobacter hormaechei subsp., K. variicola, P. mirabilis, M. morganii and Providencia spp*., are increasingly being implicated in MDR, XDR and PDR infections globally as opportunistic pathogens ^7,8,28,34,39–41^. We show herein, that *C. freundii, Enterobacter hormaechei subsp. hormaechei, xiangfangensis, oharae* and *steigerwaltii*, and Proteeae strains harbour multiple resistance mechanisms that can make them MDR, XDR and PDR pathogens. More concerning is the global distribution and multiple (human, animal, plants and environmental) specimen sources of these strains, which suggest that they can cause anthroponotic, zoonotic and food- and water-borne infections ^42^

Hence, these opportunistic pathogens demand more attention than they have been given hitherto as the rich resistome repertoire identified in their genomes makes them reservoirs of ARGs ^11,14,21,43^. Moreover, being intestinal denizens and commensals, they can easily share these ARGs with facultative and obligate pathogens of humans and animals ^22,44–46^. Further, their presence on plants and the environment further suggests that they can share their ARGs with food-borne and water-borne pathogens ^44–46^. Fortunately, the *E. xiangfangensis* strains found in rice from India had very few ARGs, albeit a few had multiple ARGs (Fig. S2B).

Of greater concern is the rich resistome repertoire and abundance of globally distributed *E. hormaechei subsp*. strains. Specifically, *E. hormaechei subsp*. contained clinically important ARGs such as *mcr-9*, carbapenemases, and ESBLs, alongside fluoroquinolones, aminoglycoside, tetracycline, macrolide, fosfomycin, chloramphenicol, rifamycin and sulphomethoxazole-trimethoprim resistance mechanisms. This resistome repertoire was also seen in *C. freundii, P. rettgeri P. mirabilis, P. stuartii*, and *M. morganii*, to a relatively lesser degree in a descending order. This is a worrying observation as colistin, carbapenems and tigecycline are last resort antibiotics used to treat fatal bacterial infections ^47,48^. In most cases, these antibiotics are used in combination with fosfomycin, fluoroquinolones and aminoglycosides to treat carbapenem-resistant Enterobacteriaceae (CRE) infections ^47,48^. Evidently, the presence of all these resistance mechanisms to these antibiotics, could make these species PDR and automatically qualify them as critical priority pathogens per the WHO criteria^32,33^.

As well, members of the tribe Proteeae viz., *M. morganii, P. mirabilis*, and *Providencia spp*., are known to have intrinsic resistance to colistin, tigecycline, aminopenicillins, amikacin, tobramycin, lincosamides, macrolides, fosfomycin and first- and second-generation cephalosporins ^1,7,49,50^. Thus, the presence of additional resistance determinants in this tribe is especially worrying. Already, there are increasing reports on the isolation of Proteeae species in recurrent urinary tract infections (UTIs) infections, which is facilitated by the increasing use of colistin to treat MDR infections; their broad intrinsic resistance mechanisms enable them to flourish during antibiotic chemotherapy ^1,7,8,49,51,52^ These observations evince the growing threat of antimicrobial resistance globally and its associated after-effects, supporting the need for efficient antibiotic stewardship to safeguard current antibiotic arsenals as well as discover novel ones ^3,14^.

As shown in the phylogenomic and phylogeographic analyses, local and international transmission, or outbreaks of strains within the same clone, clade and subclade i.e., of very close evolutionary distance, were observed. Notably, these closely related strains were isolated from humans, animals, plants, and the environment and they harboured important resistance determinants as described above. The phylogenomics showed the gradual evolution of a single strain during dissemination from host to host and depict the fact that antibiotic resistance respects no boundaries. Notably, a large part of *E. hormaechei* clade C consisted of strains from human stools/urine in Japan; these were closely related strains evolving from Spain, Taiwan, China, and Greece (Fig. 6 & S2). A similar observation was made with strains of close evolutionary distance from humans in Nigeria, France, Spain, Portugal, and Lebanon as well as with strains from humans, animals, and the environment in several countries such as France, Germany, USA, Pakistan, Morocco, Lebanon, and Poland in clade C. *E. hormaechei* strains were also found in desert sands in Morocco, showing their broad and diverse niches (Fig. 6 & S2).

Uniform and non-uniform resistome patterns were seen between strains of the same clade/clone in almost all the species. For instance, the same resistome was seen in *C. amalonaticus* clade B2 (Fig. 2), *E. cloacae* clade B (Fig. 5) and *P. mirabilis* clade C2 (Fig. 9) whilst differing resistome patterns were observed in the other species and clades. This observation supports two phenomena: firstly, the clonal expansion of strains harbouring the same resistome repertoire on both chromosomes and plasmids and secondly, the horizontal transmission of genetic elements bearing the same of different ARGs across clones and species. During clonal expansion of strains, there is a concomitant replication of resistance plasmids alongside chromosomal replication, leading to daughter cells with the same resistome diversity ^11,12,21,53^. As well, horizontal gene transfer of ARGs between bacteria can alter the resistome diversity and composition of daughter clones emanating from the same ancestor ^11,12,21,53^. In this case, both phenomena are being observed, showing that both vertical and horizontal transmission of ARGs are ongoing during the evolutionary epidemiology of the various clades and species across the globe.

The presence of multiple ARGs in a single strain might not necessarily mean they are all being expressed in the bacteria’s phenome as antibiotic-susceptible strains have been found to harbour ARGs. For instance, colistin- and fosfomocyin-sensitive Enterobacteriaceae have have been reported in strains harbouring the *mcr-9* and *fosA* genes ^1,5,7,8^. Nevertheless, the ability of these ARGs to be expressed in the presence of strong promoter or transferred to another host with a stronger promoter for subsequent expression cannot be gainsaid ^54^ Indeed, antibiotic abuse could serve as an inducer to trigger the transfer and expression of these vast resistomes in the microbial phenomes ^49,55^, necessitating the importance for judicious antibiotic use.

It is revealing to note that *K. variicola, C. amalonaticus* and *C. koseri* strains had very few ARGs except for *K. variicola* clade B2 and *C. amalonaticus* clade B2, despite the global distribution of *K. variicola* (Fig. 2-3, 7, & S3). Notably, *C. amalonaticus* clade B2, which were all from France, were remarkably enriched with ARGs including *bla*_NDM_, representing a local outbreak of XDR *C. amalonaticus* strains (Fig. 2). Thus, even in species with fewer resistome diversity and abundance, there are MDR, XDR and PDR strains causing local outbreaks.

Notably, most of the genomes included in this analysis were from the USA, Europe, and South East Asia. This may be due to the fact that these regions have higher prevalence and incidence of infections resulting from these pathogens or that these areas have enough financial and technical means to undertake genomic sequencing of these isolates in periodic surveillance studies. Specifically, genomes of these species were relatively scarce from a large part of Russia, Middle and North-West Asia, Africa, the Caribbean and parts of South America and Canada. Given the alarming resistome diversity and composition realised in this analyses, it is incumbent for all nations to intensify and adopt genome-based epidemiological studies to quickly identify the sources and reservoirs of ARGs to pre-empt outbreaks of MDR, XDR and PDR pathogens.

### Conclusion

In conclusion, less described Enterobacteriaceae species viz., *Enterobacter hormaechei subsp. hormaechei, xiangfangensis, steigerwaltii* and *oharae, C. freundii, M. morganii, P. mirabilis, P. stuartii* and *P. rettgeri*, contain globally distributed MDR, XDR and PDR strains that cause local and international outbreaks, transmitting through humans, animals, food, water and other environmental media or sources. Notably, the resistome repertoire of these relatively rare species were equally or more abundant and diverse as those of commonly isolated species. Hence, intensified efforts should be made to increase education on antibiotic stewardship to safeguard the potency of available antibiotics and reduce the selection and dissemination antibiotic-resistant Enterobacteriaceae. Infection prevention and control as well periodic genomic surveillance of communities, hospitals, farms, water bodies and the general environment (One Health) should be undertaken to pre-empt outbreaks of MDR strains and inform infection control measures.

Notwithstanding the revealing details obtained in this study, strains with clinically important ARGs whose genomes are not deposited in NCBI/PATRIC or whose genomes are not sequenced before January 2020 will be missed; hence, the information contained herein are true up to January 2020. Nevertheless, the global phylogeography and resistome epidemiology of these emerging opportunistic pathogens provide an important picture of the ARGs, sources and transmission patterns of these species.

## Methods

### Included genomes

Genomes of *Citrobacter spp*. (including *amalonaticus, freundii, koseri, werkmanii, brakii, portucalensis* and *youngae), Enterobacter hormaechei subsp. hormaechei, xiangfangensis, steigerwaltii*, and o*harae, Providencia spp*. (including *alcalifaciens, burhodogranariea, heimbachae, rettgeri, rustigianii*, and *stuartii)* and *Proteus mirabilis* deposited at GenBank (https://www.ncbi.nlm.nih.gov/genbank/) and PATRIC (https://www.patricbrc.org/) up to January 2020 were pooled and filtered to remove poor genome sequences. These were used for the downstream phylogenetics, phylogeography and resistome analyses.

### Phylogenetics and evolutionary epidemiology analyses

Briefly, plasmid sequences, phages, poor genomes i.e., genomes with sizes below the average genome size of each species viz., 3-4Mb, and genomes of strains that could not share at least 1000 core protein genes with all the included genomes were removed. The remaining genomes were aligned and run through RAXmL in batches of 200 genomes to draw phylogenetic trees using the maximum-likelihood method. A minimum of 1000 genes were used for the alignment and a bootstrap resampling of 1000x was used.The trees were annotated using Figtree to show their sequence type (ST), host (species), country and year of isolation. The various clades and sub-clades within each species or genera were visually identified based on their clustering distance and accordingly labelled.

### Phylogeography

The various clades and subclades per species were manually drawn unto maps to show their phylogeographic distribution using Paint 3D. Different colour codes were used to distinguish between the various species and clades.

### Resistome analyses

The resistomes of the included genomes were individually obtained from the NCBI Pathogen Detection database (https://www.ncbi.nlm.nih.gov/pathogens/isolates#/search/). The resistomes were aligned per strain and colour-coded per clade or species to show their association per species, clone, or clade. These were then associated with the phylogenomic trees to ascertain the resistome dynamics per clone, clade, species, and geographical location.

## Data Availability

All data are included as supplemental files

## Tweet

Global *E. hormaechei, C. freundii, P. mirabilis, P. stuartii* & *P. rettgeri* strains contain multiple resistance mechanism to important and reserved antibiotics in globally circulating clones. These portend the dawn of pandrug resistance and a return to the pre-antibiotic era.

## Highlights/Importance

*Citrobacter spp., Enterobacter hormaechei subsp., Klebsiella variicola* and *Proteae* tribe members are rarely isolated Enterobacteriaceae increasingly implicated in nosocomial infections. The global phylogenomics, evolution, geographical distribution and resistome repertoire of these species found them to be globally distributed, being isolated from human, animal, plant, and environmental sources. *E. hormaechei subsp., C. freundii, Proteus mirabilis, Providencia stuartii, Providencia rettgeri* and *Morganella morganii* contained multidrug-resistant clades that harboured resistance to clinically important reserved antibiotics, portending the dawn of pandrug resistance and a potential acceleration of the post-antibiotic era.

## Acknowledgements

None

## Transparency declaration

None (Authors declare no conflict of interest)

**Supplemental dataset 1.**
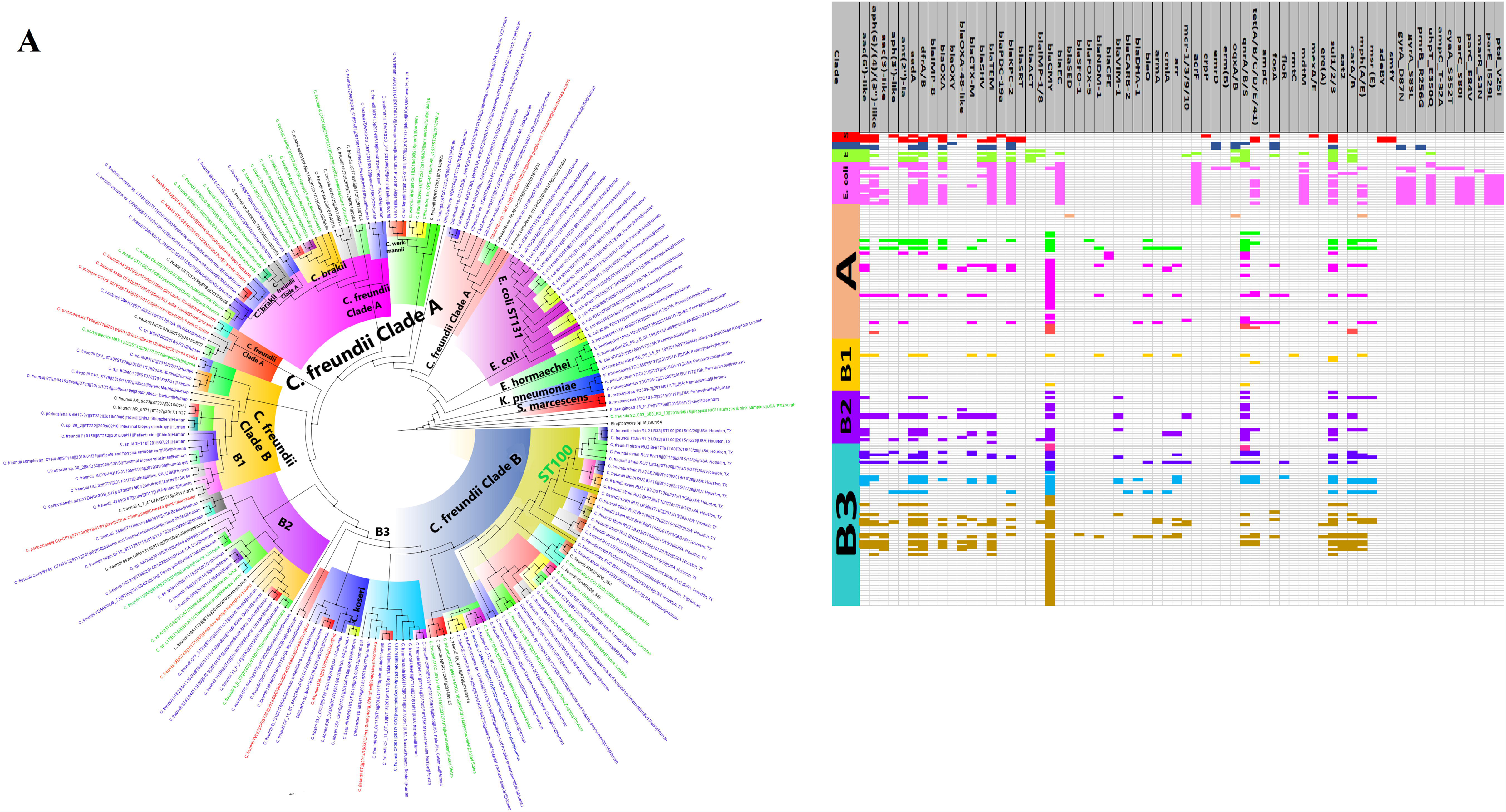
Raw metadata of downloaded genomes from PATRIC containing all the data associated with each genome.

**Supplemental dataset 2.**
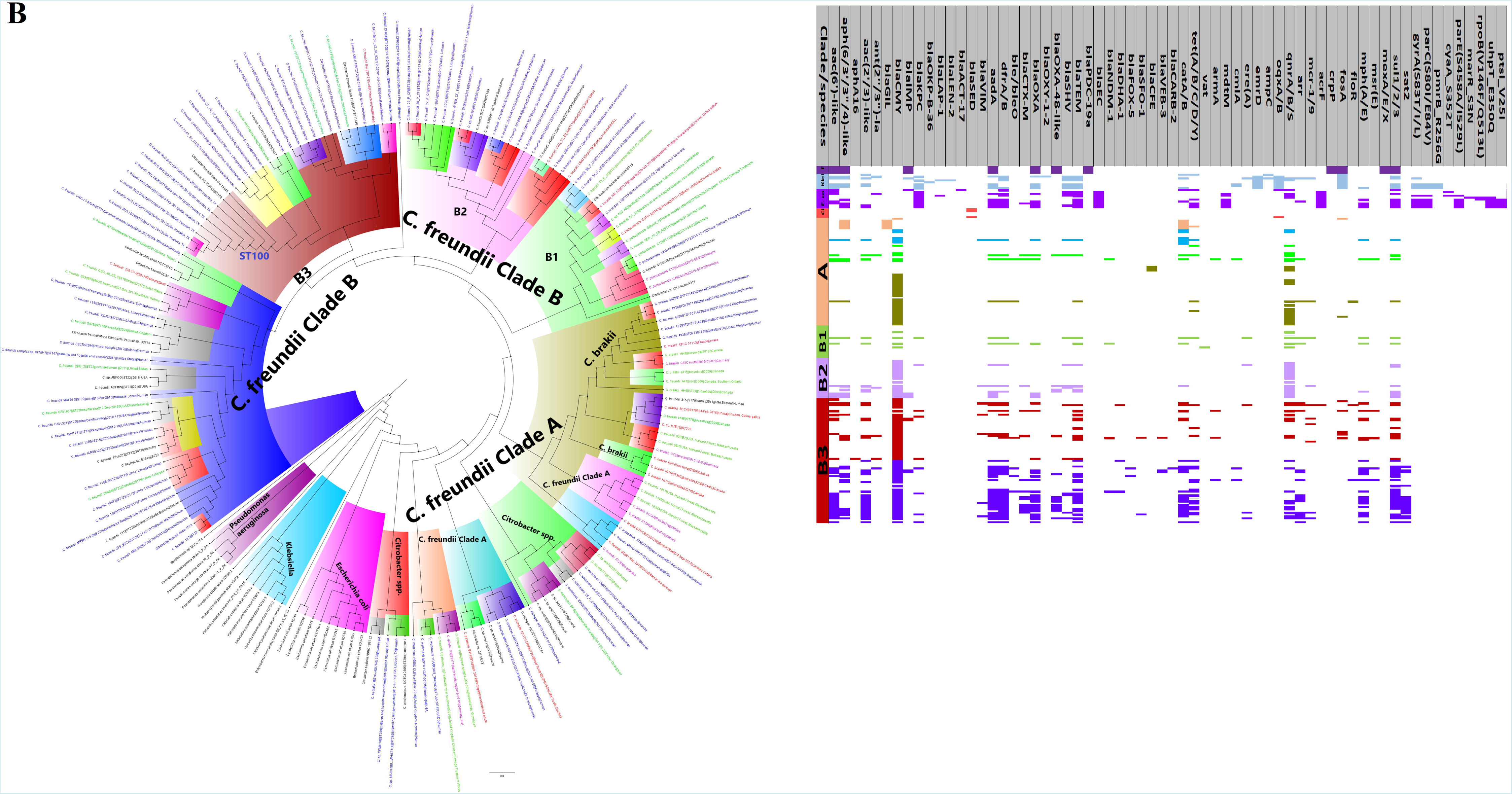
Species by species tabulation and analyses of the resistomes, specimen sources, country of isolation, MLST, Biosample accession number, and strain name of all the genomes according to their order on the phylogeny trees.

**Supplemental dataset 3.**
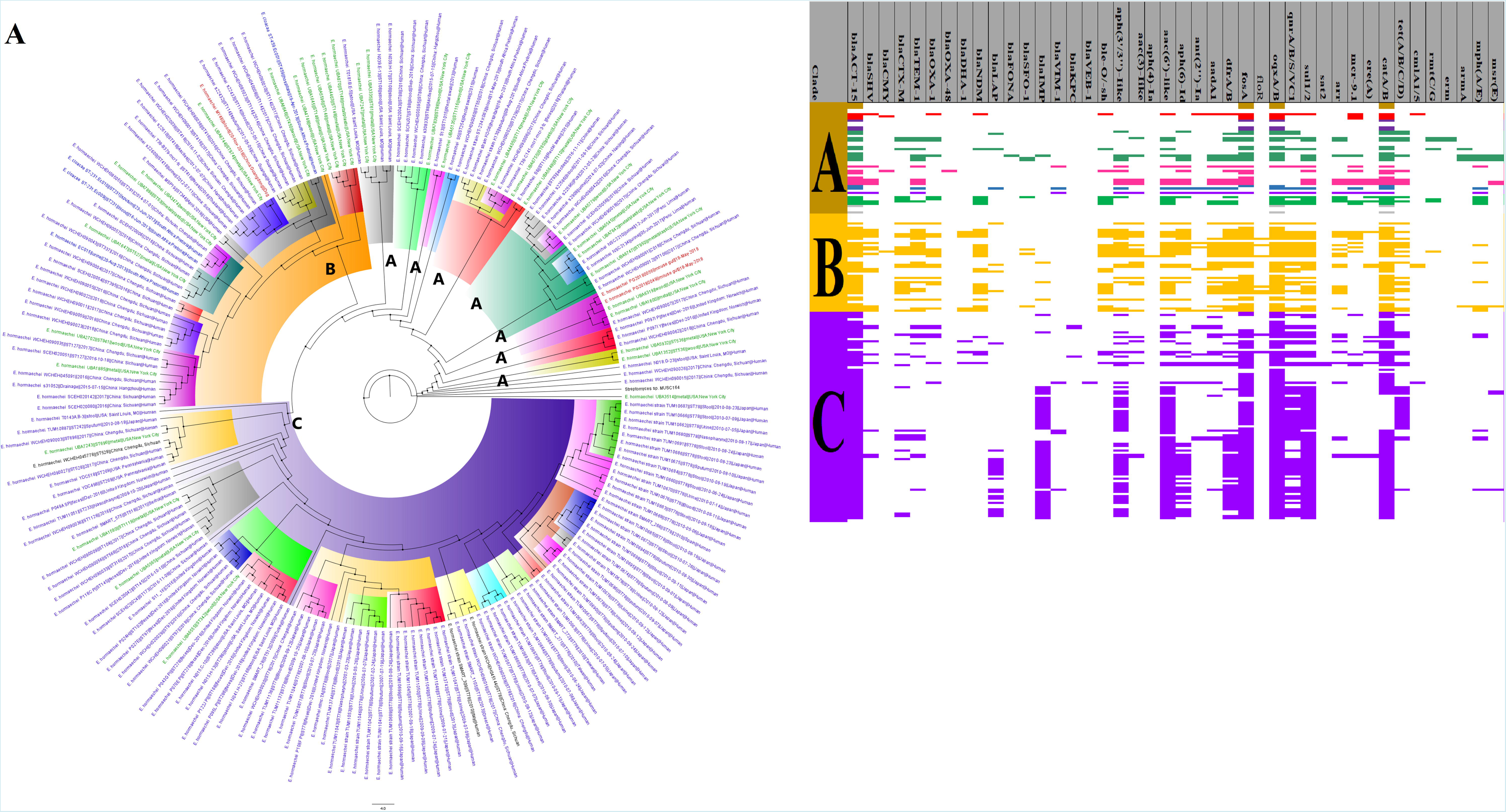
Colour-coded species by species tabulation of the resistomes, specimen sources, country of isolation, MLST, Biosample accession number, and strain name of all the genomes according to their order on the phylogeny trees.

**Figure S1 (A and B).**
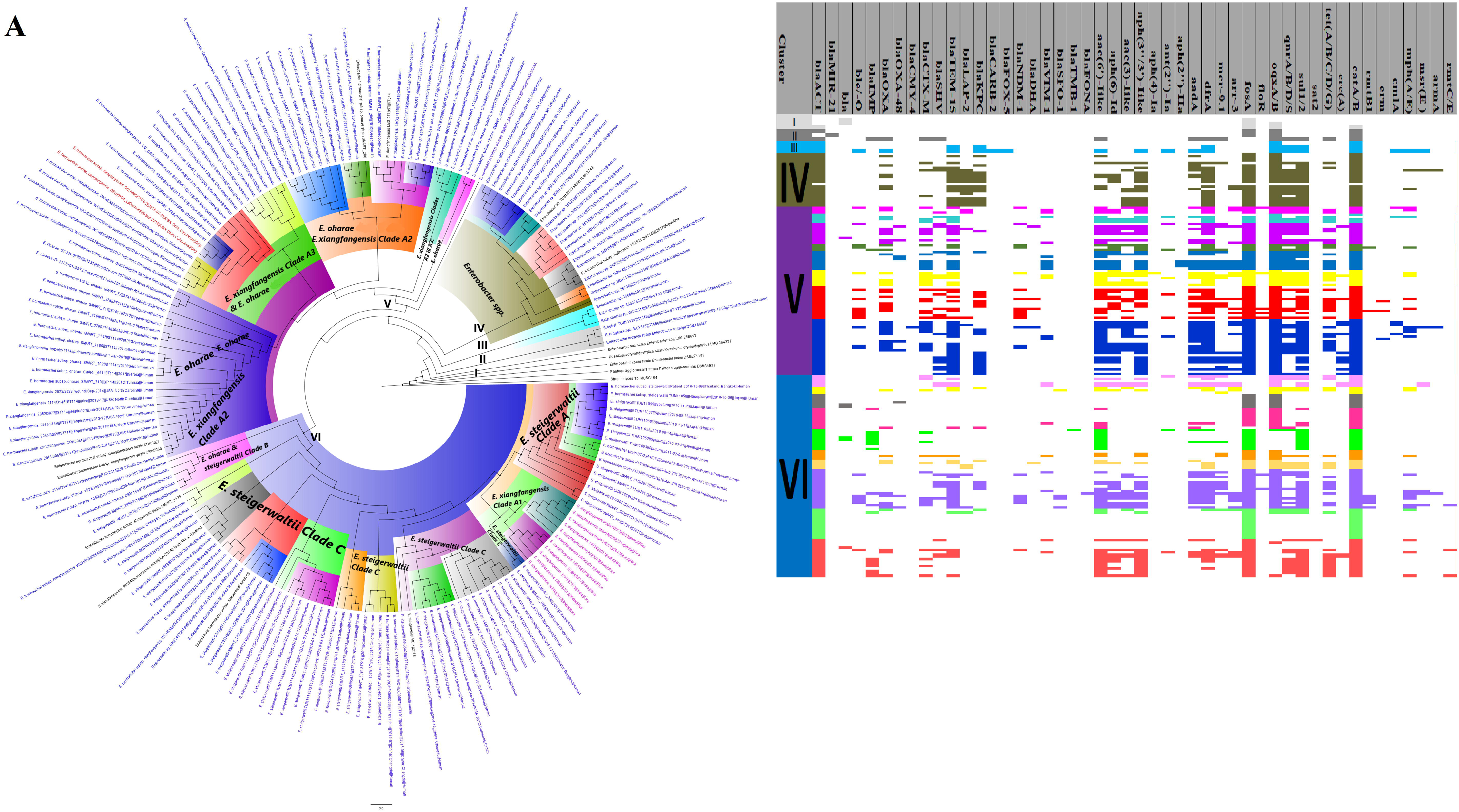
Evolutionary epidemiology and resistome of global *Citrobacter freundii* isolates, A and B. *C. freundii* clustered into four main clades (A, B1, B2 and B3), highlighted with distinct colours. Clade B3 had the most resistome abundance and diversity. Strains from humans (blue labels), animals (red labels), plants (purple/mauve labels) and the environment (green labels) were found in the same clade/cluster. *Bla*_CMY_ was conserved in these genomes.

**Figure S2 (A and B).**
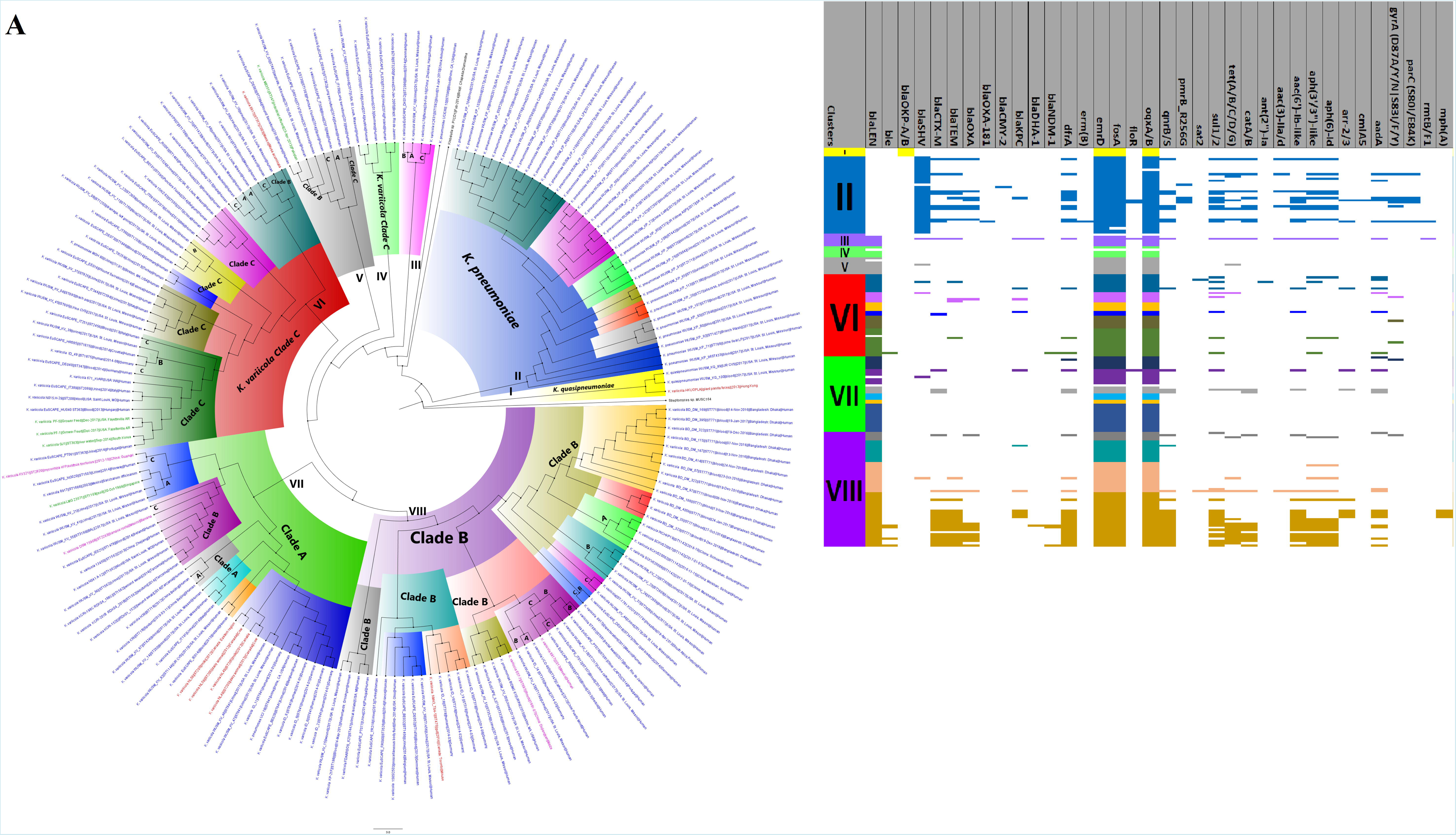
Evolutionary epidemiology and resistome of global *Enterobacter hormaechei* isolates, A and B. The *E. hormaechei* isolates clustered into three main clades A, B and C (with distinct highlights) that contained strains distributed globally from humans (blue labels), and animals (red labels), plants (purple/mauve labels) and the environment (green labels). Clades B and C contained diverse and rich resistome repertoire. *bla*_ACT_ was conserved in these genomes.

**Figure S3 (A and B).**
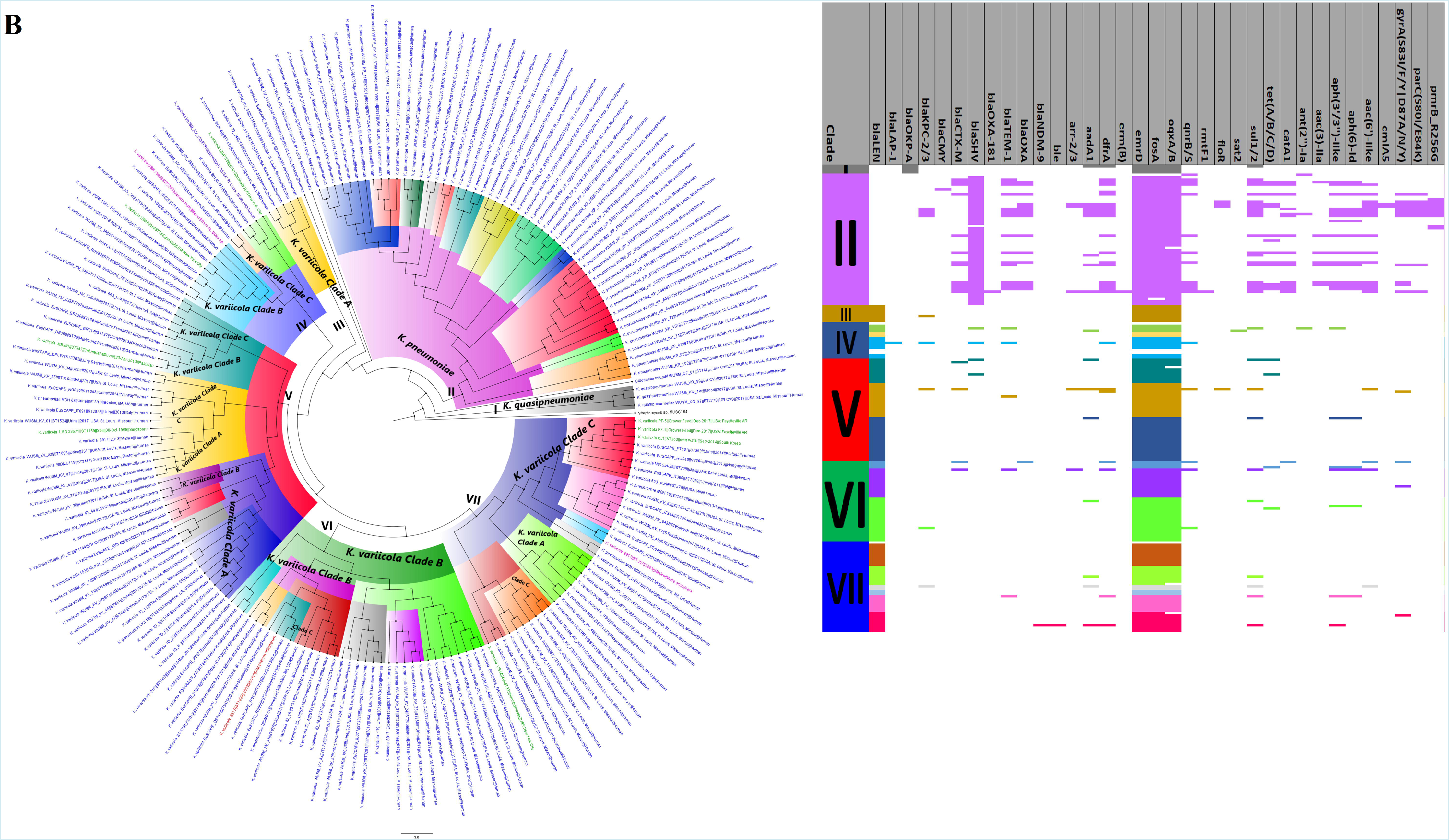
Evolutionary epidemiology and resistome of global *Klebsiella variicola* isolates, A and B. The *K. variicola* strains clustered into nine clades viz., A1, A2, A3, A4, A5, A6, B1, B2 and C, which were highlighted with distinct colours and were isolated from countries around the globe. The clades contained strains distributed globally from humans (blue labels), animals (red labels), plants (purple/mauve labels) and the environment (green labels). Besides a few strains in clade B2, the other strains contained very few resistance genes. *bla*_LEN_ was conserved in these genomes.

**Figure S4.**
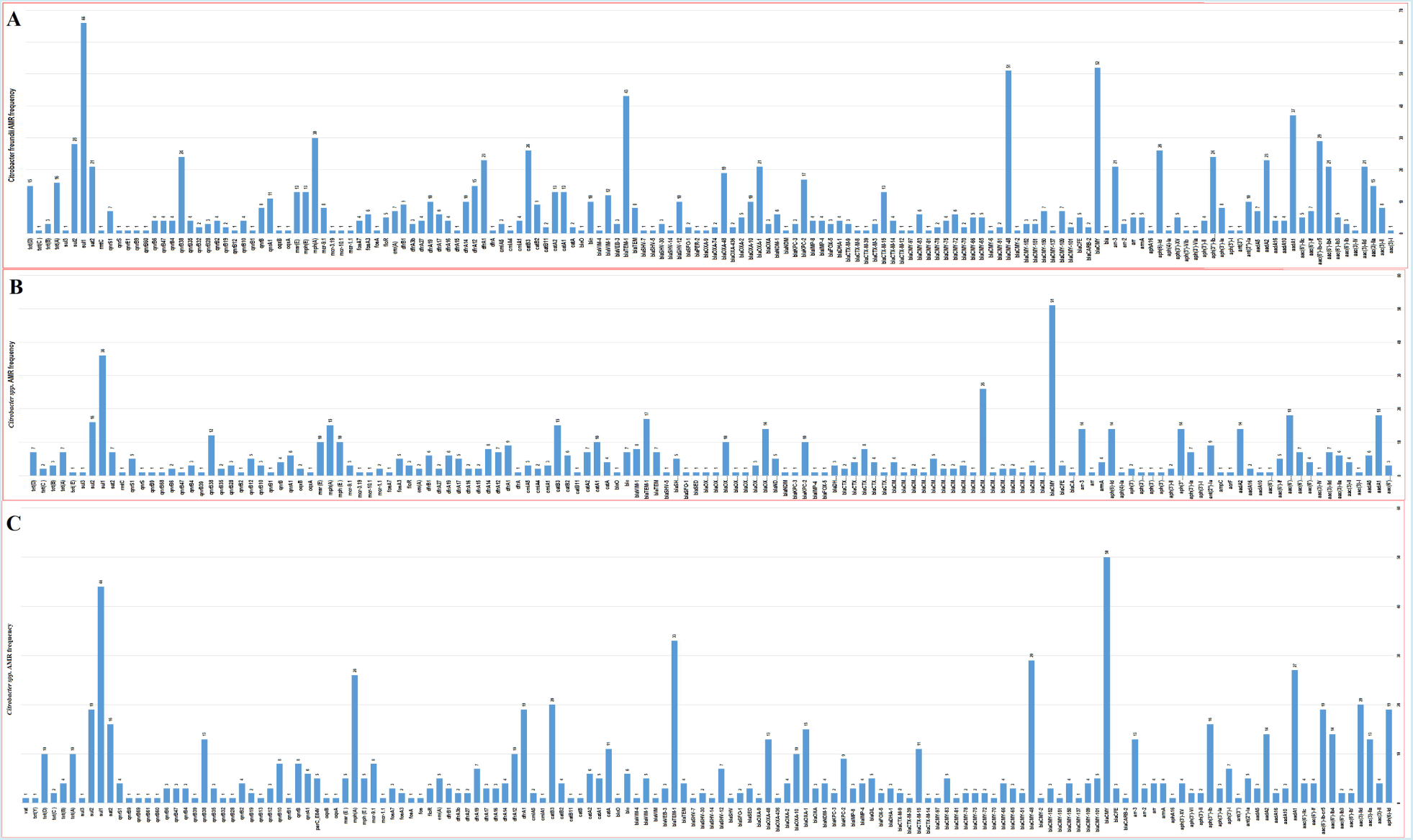
Frequency distribution of antibiotic resistance genes in *Citrobacter freundii* (A), and *Citrobacter species* (B and C).

**Figure S5.**
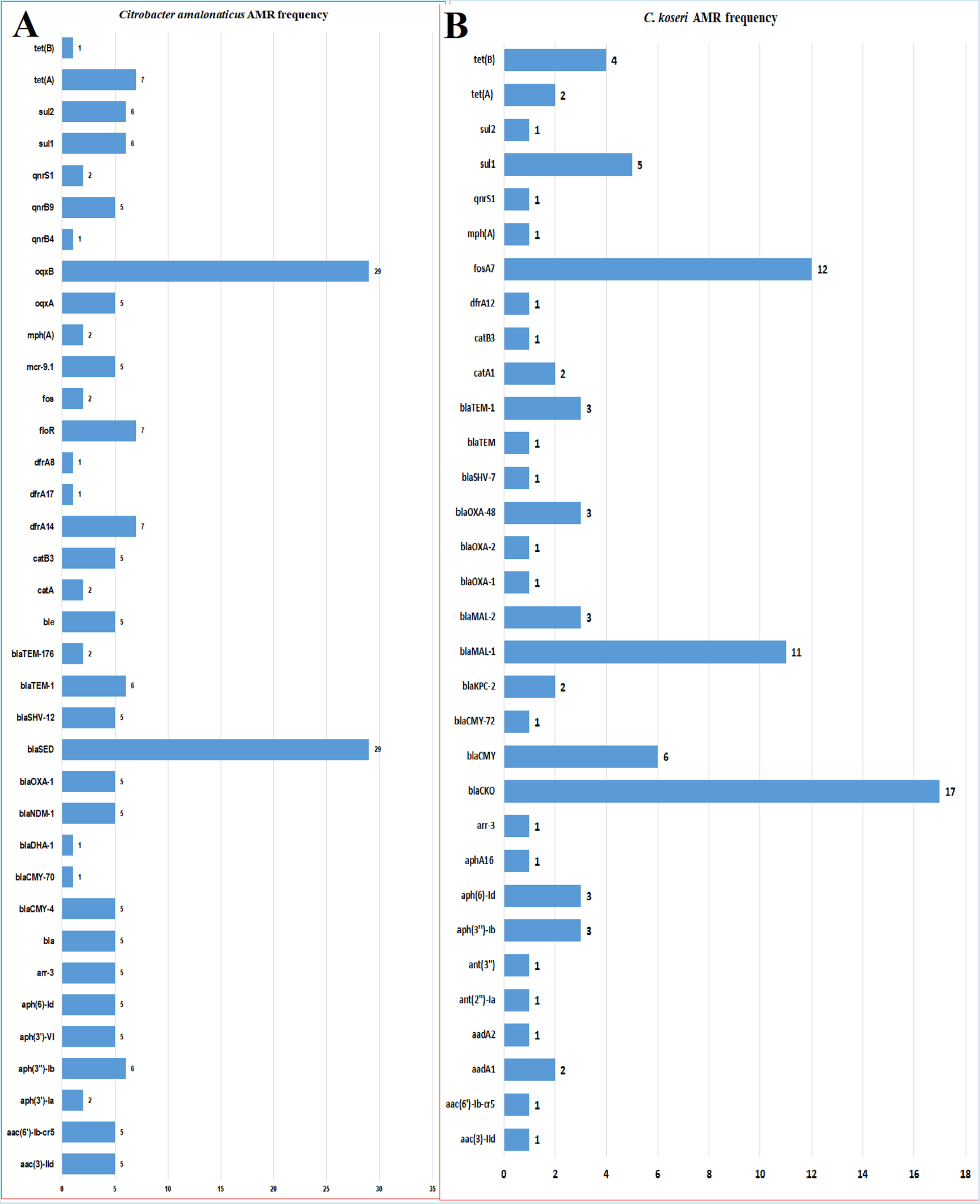
Frequency distribution of antibiotic resistance genes in *Citrobacter amalonaticus* (A), and *Citrobacter koseri* (B).

**Figure S6.**
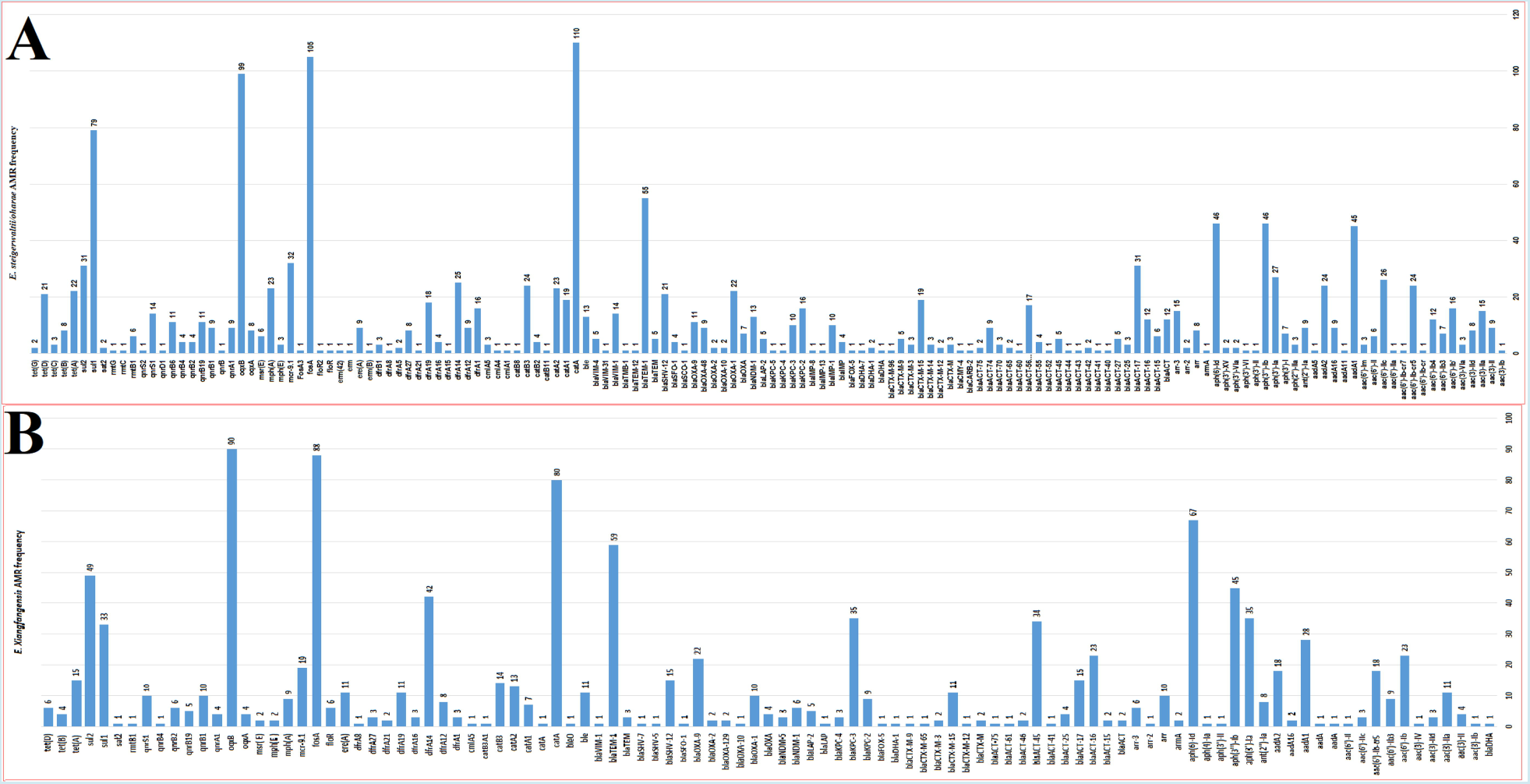
Frequency distribution of antibiotic resistance genes in *Enterobacter steigerwaltii* and *oharae* (A), and *Enterobacter xiangfangensis* (B).

**Figure S7.**
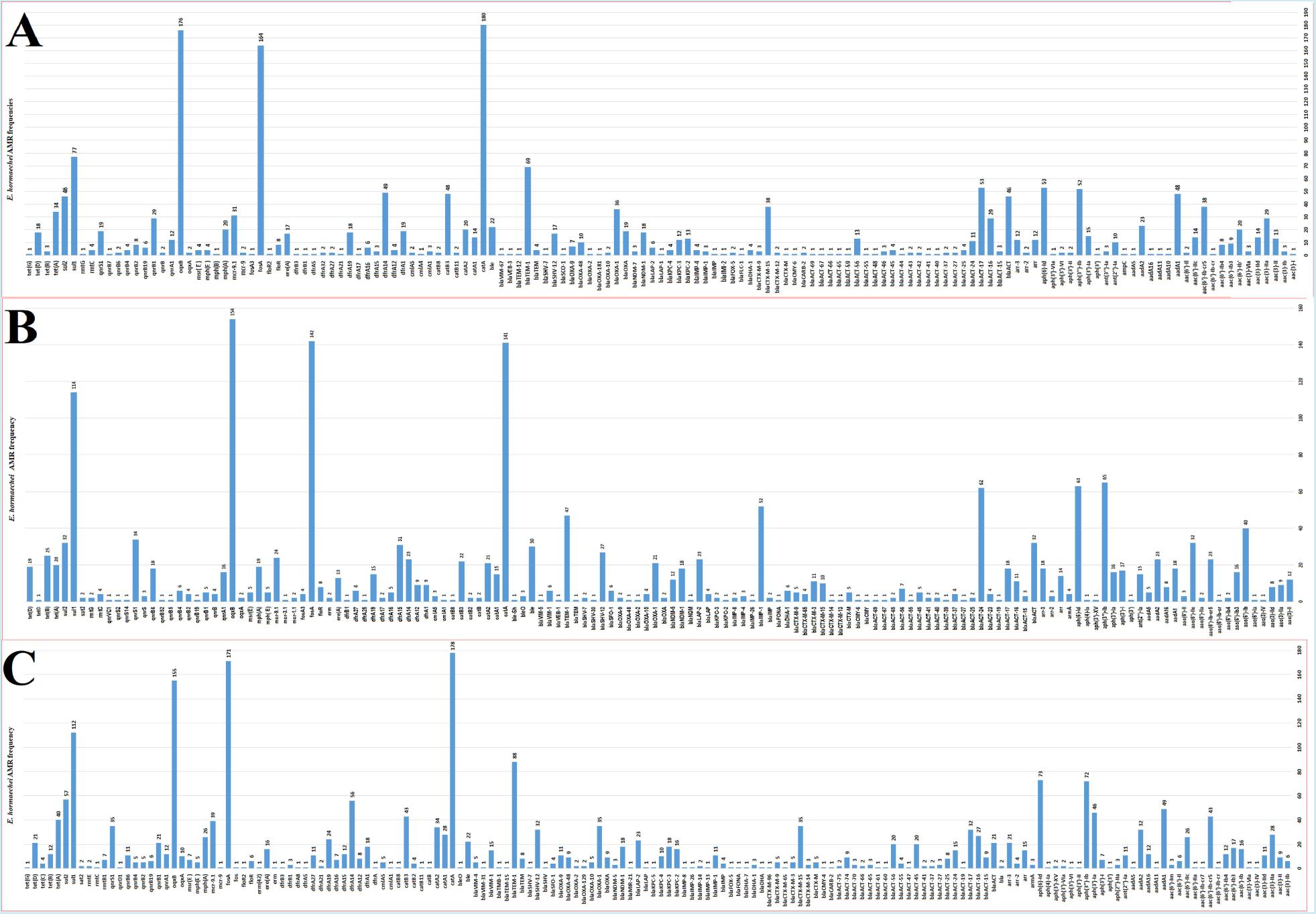
Frequency distribution of antibiotic resistance genes in *Enterobacter hormaechei* (A, B and C).

**Figure S8.**
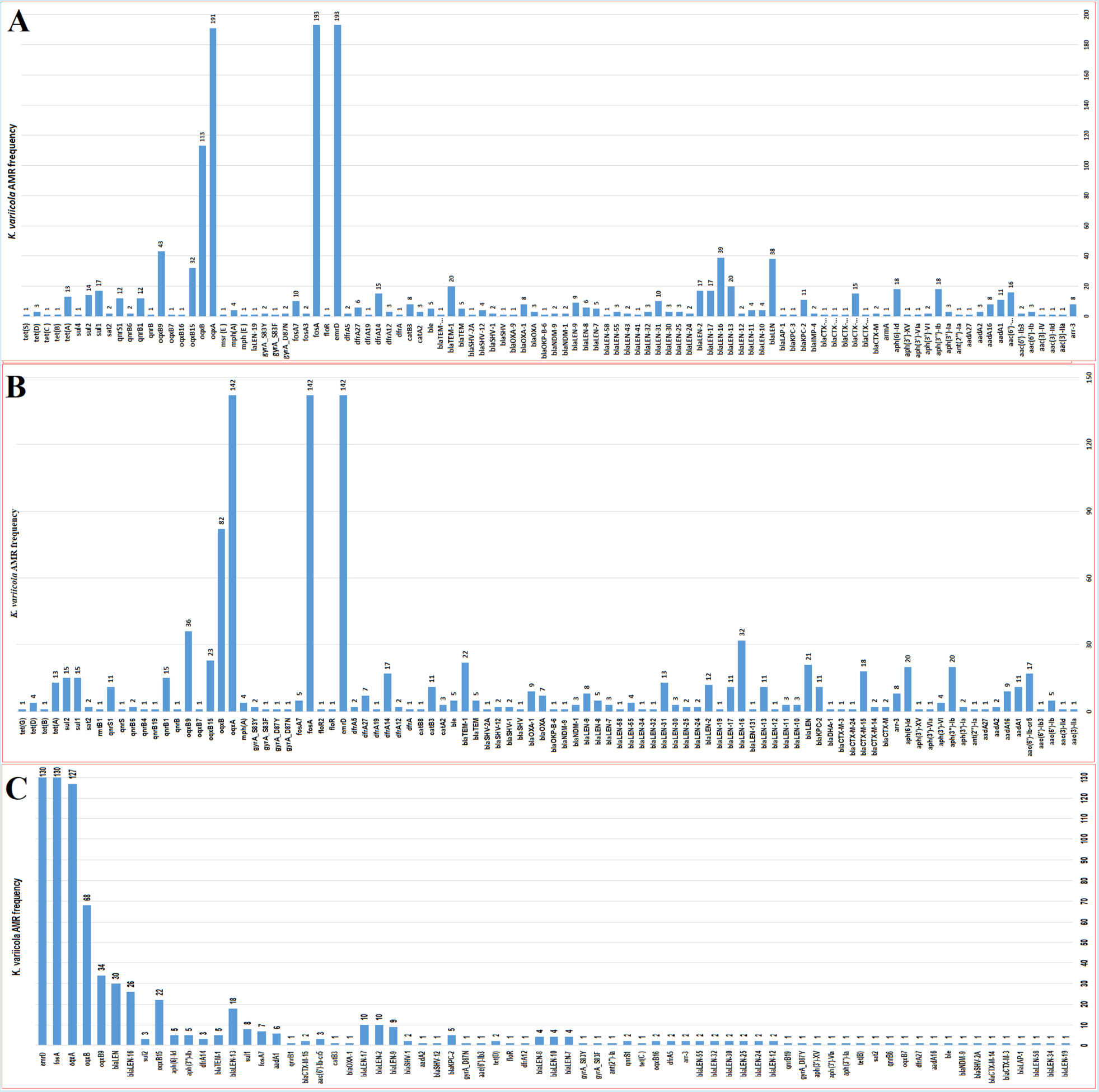
Frequency distribution of antibiotic resistance genes in *Klebsiella variicola* (A, B and C).

**Figure S9.**
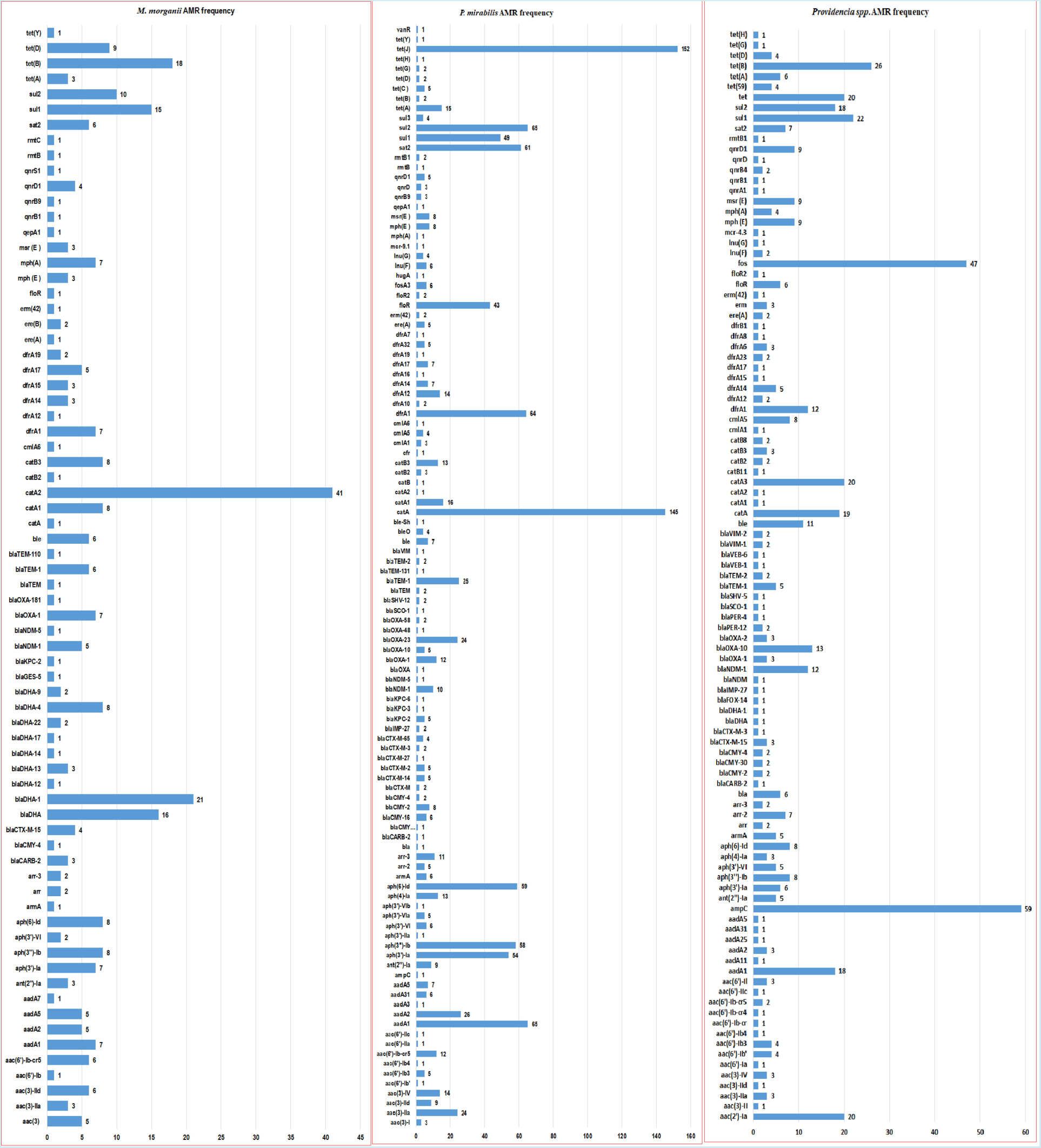
Frequency distribution of antibiotic resistance genes in *Morganella morganii* (A), *Proteus mirabilis* (B) and *Providencia species*(C).

